# Wellbeing While Waiting: Effectiveness and implementation of youth social prescribing for young people awaiting CAMHS support

**DOI:** 10.64898/2026.03.11.26348130

**Authors:** D. Hayes, J. Wright, A. Burton, F. Bu, L. Sticpewich, H. Stuttard, J. Page, A. Bradbury, E. Han, J. Deighton, M.S. Tibber, S. Talwar, D. Fancourt

## Abstract

**Background:** Prolonged waiting times for Child and Adolescent Mental Health Services (CAMHS) leave many young people without structured support while awaiting specialist treatment. Social prescribing has been proposed as a community-based adjunct within CAMHS pathways; however, evidence regarding its safety and clinical impact remains limited.

**Methods:** Wellbeing While Waiting was a multi-site non-randomised controlled trial embedded within a hybrid type II implementation–effectiveness evaluation conducted across 11 CAMHS in England. The protocol was prospectively published prior to recruitment (BMC Psychiatry; 10.1186/s12888-023-04758-0). Between May 2023 and March 2025, 558 young people aged 11–18 years referred to CAMHS were enrolled (225 usual care; 333 social prescribing). Primary outcomes were anxiety and depression symptoms, total emotional and behavioural difficulties, and perceived stress. Secondary outcomes included resilience and wellbeing.

**Results:** No intervention-related adverse events were observed. On average, participants had 5 sessions with a Link Worker. Compared with usual care, no significant differences were observed in anxiety or depression symptoms. However, participants receiving social prescribing demonstrated significant improvements in total emotional and behavioural difficulties over six months, driven by reductions in conduct difficulties, hyperactivity and peer problems. Significant improvements for those receiving social prescribing were also found for prosocial behaviour and resilience.

**Conclusions:** Within routine CAMHS pathways, no intervention-related adverse events were observed for social prescribing, and social prescribing was associated with improvements in behavioural and resilience-related outcomes, although not in anxiety or depressive symptoms. Findings suggest social prescribing may offer a valuable adjunct during delayed access to specialist treatment, with effects distinct from symptom-focused clinical therapies.

## Introduction

Child and youth mental ill health is a growing concern. Over the last decade, many countries have been reporting increases in prevalence rates, with sharp rises during the pandemic [1]. Globally, amongst children and young people, anxiety disorders are now the leading cause of non-fatal disability, and depressive disorders are placed fourth [1]. Within the UK, latest estimates indicate that 1 in 5 young people now have a probable mental health disorder [2]. As a result, child and adolescent mental health services (CAMHS) face increased demand from young people and their families for mental health support [3]. National data indicate that as of late 2024, over 350,000 children and young people were waiting for specialist mental health care in England alone, with median wait times of just under eight months, and some waiting more than a year for treatment [4]. These far exceed the government’s proposed policy target of four weeks [5] and are disproportionately experienced by young people living in socioeconomically deprived areas, where need is higher and service capacity more constrained [6].

Evidence increasingly suggests that waiting for mental health treatment is an important period. Qualitative and quantitative research shows that many young people experience deterioration in mental and physical health while on waiting lists, alongside increased distress, maladaptive coping strategies, and heightened risk of crisis presentations [7–10]. Families often report significant strain during this period, with caregivers attempting to compensate for the absence of professional support while navigating unclear referral pathways and limited communication from services [7]. Recent qualitative evidence demonstrates that deterioration is not solely attributable to the passage of time but is often directly linked to the experience of being placed on a waiting list, characterised by uncertainty, perceived rejection, and a mismatch between need and available support [7, 9, 10].

Despite the scale and consequences of waiting lists, interim support options for young people awaiting CAMHS treatment remain under-researched. A recent review of waiting-list interventions for CAMHS identified 18 different interventions, many of which focused on psychoeducation alongside other components, such as parental support [11]. Findings indicated that most interventions were under five sessions and aimed at parents/guardians but showed evidence of promise in improving child and youth clinical outcomes and/or were seen as feasible/acceptable [11]. However, caution in findings was advised due to the overall low methodological quality of papers, including small sample sizes and a lack of control groups. As a result, the review called for more youth-co-designed, pragmatic trials and service evaluations to evaluate real-world impact of further interventions.

Social prescribing (SP) is a mechanism of care that connects individuals to non-clinical, community-based activities and resources intended to support health and wellbeing [12]. This process usually involves a health or social care professional referring a patient to a Social Prescriber, sometimes known as a Link Worker (LW), who develops a non-clinical plan that connects the patient with community organisations to improve health, wellbeing or other aspects of the patient’s life [13]. Activities include the arts, cultural events, and other support services, such as physical activity, financial support, volunteering and befriending [12]. Within the English National Health Service (NHS), SP has been embedded within universal personalised care and expanded rapidly in adult primary care settings [14]. For adults, SP has been associated with improvements in mental health, wellbeing, quality of life, and social connectedness, as well as reductions in loneliness and service use [15–20]. However, the extension of SP to children and young people has lagged behind adult provision [21]. Whilst less developed, the growing evidence base suggests it can have similar benefits for young people, with ‘evidence of promise’ across areas such as mental health, wellbeing and loneliness [22, 23].

With few evidence-based child- and youth-focused support available during long waits for CAMHS treatment, SP is increasingly being adopted as a waiting-list intervention. It aims to address social factors that may precipitate or perpetuate distress, offers a non-pharmacological route whilst young people wait for support, facilitates engagement in community-based activities that may reduce stigma and “medicalisation”, and aligns with personalised care by increasing choice and control over patients’ support [24]. As SP is increasingly piloted within CAMHS as a waiting-list intervention, Wellbeing While Waiting was designed to evaluate both clinical impact and real-world implementation, using routine quantitative delivery indicators (e.g., reach and dose) and brief acceptability/feasibility measures alongside outcome assessment. This paper reports the main quantitative implementation and effectiveness findings from a pragmatic real-world controlled evaluation, addressing a key evidence gap in youth SP: whether it improves mental health outcomes for young people awaiting CAMHS treatment.

## Methods

### Design

Wellbeing While Waiting was a multi-site non-randomised controlled trial embedded within a hybrid type II implementation–effectiveness evaluation of a co-designed youth SP pathway delivered across 11 CAMHS services in England [25]. The pathway was developed with input from young people through patient and public involvement and engagement activities via a Youth Advisory Group (YAG) that informed the intervention model and study materials. The YAG also helped interpret findings. The primary aim of the quantitative evaluation was to assess both the effectiveness of SP on mental health and wellbeing outcomes for young people awaiting CAMHS treatment, and key implementation outcomes captured through routine service data. Implementation was quantified using delivery metrics including reach and intervention dose, alongside brief questionnaire measures of acceptability and feasibility; qualitative findings on Wellbeing While Waiting from stakeholders are reported separately [26].

Random allocation was not feasible due to the staged roll-out of the SP pathway across participating sites. Sites initiated recruitment to the usual care (pre-implementation) cohort between June 2023 and March 2024 and subsequently began recruiting to the SP (post-implementation) cohort at different time points between June 2023 and November 2024, once local implementation arrangements were in place. Site-level implementation dates are presented in the supplementary material (S1). As such, this study adopted a pragmatic non-randomised stepped wedge design, comparing outcomes for young people referred to SP with those of a control group recruited prior to pathway implementation at the same sites, providing a clear temporal counterfactual. A summary of the design is provided here, and full details and the rationale for the design choice are given in the supplementary materials (S2). Quantitative data were collected at baseline, three months, and six months.

This study was conducted in accordance with the ethical standards of the institutional and/or national research committee and in line with the 1964 Helsinki Declaration and its later amendments, or comparable ethical standards. Ethical approval was obtained from the West of Scotland Research Ethics Committee 5 (Reference: 22/WS/0184). The protocol was prospectively published prior to recruitment [25]. The design, conduct, and reporting of the study followed CONSORT 2010 guidance [27], with the extension for pragmatic trials [28]. Written informed consent and, where appropriate, assent were obtained from all individual participants included in the study. For participants under the age of 16, written informed consent was also obtained from parents or legal guardians prior to participation. The CONSORT checklists are shown in the supplementary materials (S3).

### Setting

The study was conducted across 11 CAMHS sites located in England, selected to ensure variation in geography, population characteristics, and service configuration. One site withdrew during usual care recruitment due to service-level pressures (Site 5). Participating sites were planning to implement a youth SP pathway during the study period. All sites were specialist outpatient CAMHS and were experiencing extended waiting times of at least three months for evidence-based psychological treatments.

Sites differed in local voluntary and community sector infrastructure, existing partnerships and workforce configuration, but all delivered the intervention within a shared overarching model and adhered to common research procedures. SP was delivered either by LWs embedded within CAMHS teams (Sites 2 and 11) or by LWs hosted externally in partner organisations and linked to CAMHS through formal referral arrangements (Sites 1, 3, 4, 6–10). Two sites delivered sessions remotely only (Sites 3 and 8), and one site did not commence SP delivery during the study period (Site 5). Further site-level details are provided in the supplementary material (S4).

### Participants

Participants were children and young people aged 11–18 years who had been referred to CAMHS for a short-term evidence-based psychological intervention and had been on the waiting list for less than one month at the point of recruitment. Young people aged 16–18 years provided written informed consent; those aged 11–15 years provided assent alongside written parental or caregiver consent. Exclusion criteria were applied to ensure clinical appropriateness and safety. Young people were excluded if they presented with eating disorders, psychosis, or severe and complex mental health difficulties, as judged by CAMHS clinicians; these presentations typically require alternative care pathways and more rapid access to specialist treatment. Participation in the study did not delay or replace any clinical care.

### SP (active support during the waiting period)

The intervention comprised a structured, youth-focused SP pathway delivered by trained LWs or equivalent roles (e.g., community connectors) as an active support offered during the CAMHS waiting period. The pathway was person-centred and flexible. LWs met individually with young people, typically for around six sessions, although the number, frequency, duration and mode of delivery (in-person, online or telephone) were tailored to individual needs and preferences, where possible. Sessions focused on understanding “what matters” to the young person, identifying strengths and goals, and supporting access to relevant community-based activities and resources (e.g., arts programmes, sports, peer support groups, or practical support services). Young people could access a personalised budget of up to £40 to reduce financial barriers to participation (e.g., for travel, equipment or activity fees), administered by the LW. Caregiver involvement was flexible and guided by young people’s preferences and clinical appropriateness.

### Counterfactual (usual waitlist)

Young people in the control group received usual care while on the CAMHS waiting list, following referral and triage. This typically consisted of waiting-list management and standard administrative processes, with limited structured interim support. Depending on local practice, young people may have received signposting to self-help resources and/or crisis contact information. However, they did not receive a structured programme of link-worker support, facilitated community engagement, or access to a personalised budget during the waiting period.

### Measures

#### Young people

Outcome measures (baseline, three months, and six months) using validated self-report instruments.

#### Primary outcomes

Three primary outcomes focused on youth mental health and included:

- Emotional and behavioural difficulties measured using the self-report Strengths and Difficulties Questionnaire (SDQ) [29].
- Anxiety and depression symptoms measured using the Revised Child Anxiety and Depression Scale (RCADS) [30].
- Perceived stress measured using the Perceived Stress Scale (PSS) [31].

#### Secondary outcomes

- Resilience assessed using selected subscales from the Student Resilience Survey (SRS) [32].
- Subjective wellbeing assessed using Office for National Statistics (ONS) wellbeing items [33].
- Acceptability of Intervention Measure (AIM) [34].

#### LWs and CAMHS staff

Implementation measures were administered at least 6 months after SP had been implemented in the service using validated self-report instruments:

- Acceptability of Intervention Measure (AIM) [34].
- Feasibility of Intervention Measure (FIM) [34].
- Intervention Appropriateness Measure (IAM) [34].

Routine outcome data from LW host organisations was also drawn upon. This included

- The number of sessions they worked with the young person
- Any activities the young person was referred to, and
- Whether their personalised care budget was used.

### Procedure

Eligible young people were identified by CAMHS clinicians at participating sites. For the control group, recruitment occurred prior to SP pathway implementation and whilst pathways were being developed. For the intervention group, recruitment occurred after SP pathways were live at each site. Upon expressing an interest after being put on the CAMHS waiting list, participants received an age-appropriate Participant Information Sheet and were given the opportunity to ask any questions. Those who were happy to take part provided informed consent or assent.

Baseline questionnaires were completed at enrolment, with follow-up assessments conducted at three and six months. Questionnaires were completed online, in person, or over the phone/teams, depending on participant preference. All participants received vouchers of £10 at each assessment point to acknowledge their time for participating. Research staff at NHS Trust site R&D departments and Research Assistants at UCL maintained contact with participants to support retention and minimise missing data. With assent/consent, routine service data were accessed from CAMHS/LW host organisations (if separate from CAMHS) to obtain further information on SP sessions. Owing to a questionnaire formatting error, responses for the youth AIM [34] were captured on a four-point scale (rather than five) with ‘completely agree’ being omitted. This meant that responses ranged from one (completely disagree) to four (agree).

### Statistical analysis

Data were analysed using growth curve modelling to depict changes in each of the outcome measures across three time points (baseline, three-month and six-month follow-up). Time was modelled as a discrete variable, allowing for non-linear trajectories. In addition to intervention group, predictors in each model included age (centred at age 11), gender, ethnicity, free school meals, and IMD quintiles to account for potential differences at baseline. Moreover, we included an interaction term between intervention group and time variable to assess differences in growth rate by intervention groups. We allowed intercept to vary across participants. Time, however, was treated as fixed across participants due to a small number of time points. The original published study protocol had proposed using difference-in-difference analysis, but growth curve modelling was adopted instead when the full statistical analysis plan was drawn up, as this statistical approach has greater efficiency (e.g. better handling of missing data and using all available information) and allows richer description of changes across time points and between groups. This change to the statistical analysis plan was made prior to any data being analysed. All analyses were conducted in Stata V18.0. A p-value < 0.05 was considered statistically significant, and p-value < 0.1 as marginally significant.

## Results

Between May 2023 and March 2025, a total of 558 participants were enrolled in the study, including 225 in the control group and 333 in the intervention group. Figure 1 shows the flow of participants through the study from enrolment to follow-up.

**Figure 1:**
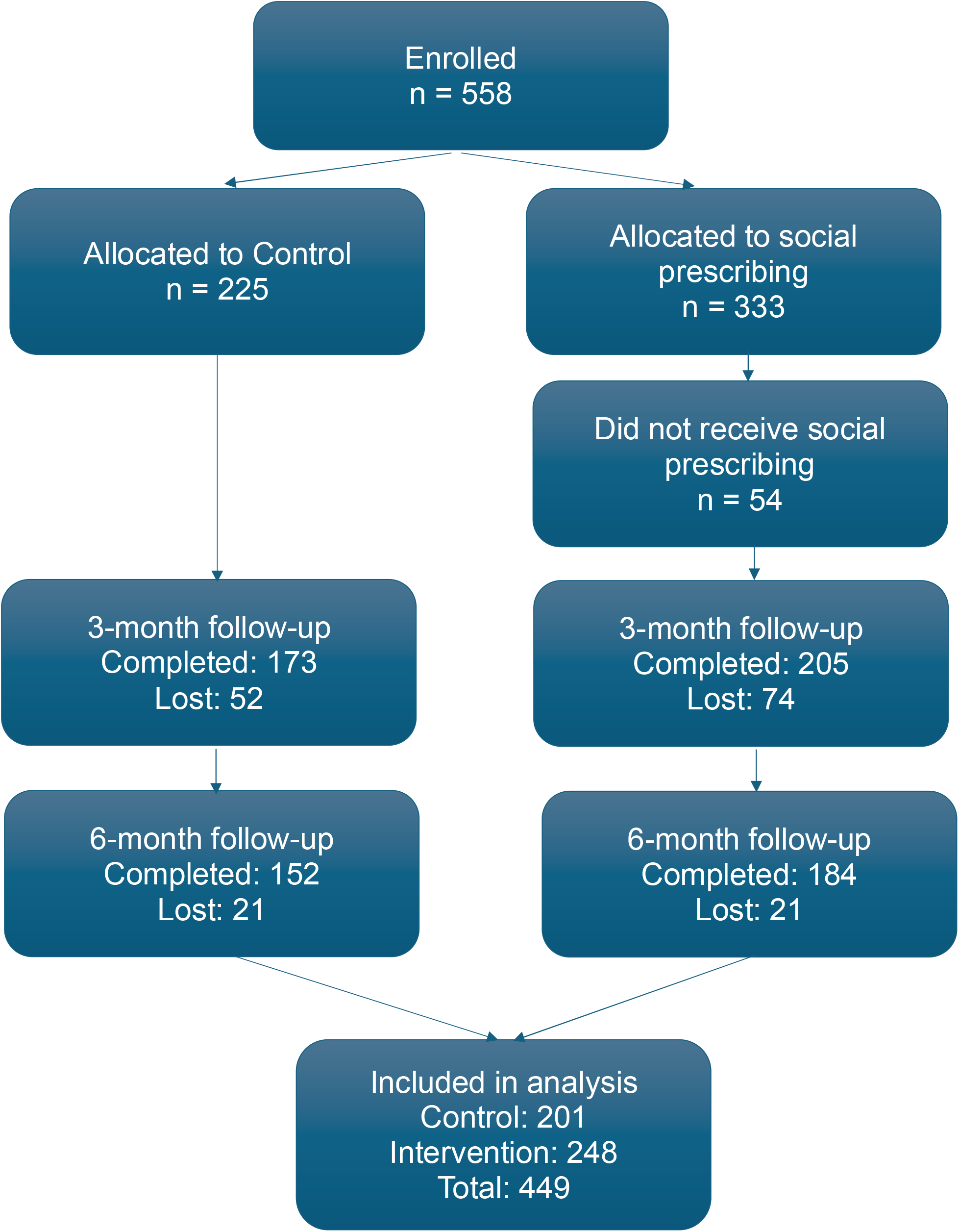
CONSORT flow diagram of participant recruitment, allocation, follow-up and analysis

### Implementation

Of the young people in the SP group (n=333), 54 participants (16.2%) did not receive SP (e.g. due to lack of parental consent, non-contact). Among the remaining participants, 231 had valid information on LW sessions, with a mean of 5.05 sessions attended (standard deviation (SD) of 2.56).

A total of 193 young people (69.2% of those who received SP) completed the Acceptability of Intervention Measure at the three-month follow-up. The majority of participants reported neutral or positive attitudes toward each item, with only a small proportion expressing disagreement (10.4–14%)

In total, 25 CAMHS clinicians and 20 LWs completed information on feasibility, acceptability and appropriateness [34]. Of these, 75.6% identified as female, 13.3% as male and 11.1% as non-binary or did not report their gender. The majority of participants (80.0%) were from a white ethnic background, and 20.0% from an ethnic minority background or did not report. Overall, 40/45 (89%) participants completed the acceptability measures, yielding a mean score of 4.5 (SD=0.61) on a five-point scale from one (completely disagree) to five (completely agree). The appropriateness and feasibility measures were each completed by 39 participants. On the same five-point scale, the mean score for appropriateness was 4.23 (SD = 0.80), and the mean score for feasibility was 4.34 (SD = 0.63). The distribution of participants responses regarding feasibility, acceptability and appropriateness are outlined in Supplementary Material S5). There was little difference between CAMHS clinicians and LWs in their ratings of the acceptability, appropriateness, and feasibility of SP.

During the study, 15 adverse events were reported and reviewed by the Data Monitoring Committee. The most common reason was a deterioration in symptoms (n=9). No adverse events were deemed related to the SP pathway.

### Effectiveness

For analyses of effectiveness, we excluded the 54 participants who did not receive SP based on LW records, and excluded a further 55 participants with missing data in baseline socio-demographic characteristics (10.9%), providing an analytical sample of 448 participants, including 201 in the control group and 248 in the intervention group (see Figure 1). Table 1 outlines the sociodemographic characteristics of participants by intervention group.

**Table 1.** Sample characteristics by intervention groups.

|  | Total<br>(n=449) |  | Control group<br>(n=201) |  | Intervention (SP) group<br>(n=248) |  |
| --- | --- | --- | --- | --- | --- | --- |
|  | Freq | Mean (SD) / % | Freq | Mean (SD) / % | Freq | Mean (SD) / % |
| Age | 449 | 13.97 (1.72) | 201 | 13.94 (1.79) | 248 | 14.00 (1.68) |
| Gender: male | 140 | 31.2% | 59 | 29.4% | 81 | 32.7% |
| Gender: female | 298 | 66.4% | 134 | 66.7% | 164 | 66.1% |
| Gender: other | 11 | 2.4% | <10 | 4.0% | <5 | 1.2% |
| Ethnicity: white | 369 | 82.2% | 157 | 78.1% | 212 | 85.5% |
| Ethnicity: other | 80 | 17.8% | 44 | 21.9% | 36 | 14.5% |
| School meals: yes | 151 | 33.6% | 68 | 33.8% | 83 | 33.5% |
| School meals: no | 298 | 66.4% | 133 | 66.2% | 165 | 66.5% |
| IMD 1 (most deprived) | 139 | 31.0% | 51 | 25.4% | 88 | 35.5% |
| IMD 2 | 108 | 24.1% | 55 | 27.4% | 53 | 21.4% |
| IMD 3 | 91 | 20.3% | 41 | 20.4% | 50 | 20.2% |
| IMD 4 | 73 | 16.3% | 36 | 17.9% | 37 | 14.9% |
| IMD 5 (least deprived) | 38 | 8.5% | 18 | 9.0% | 20 | 8.1% |

The mean age of the analytical sample was approximately 14 years (SD=1.72). Two thirds of participants identified as female, 31.2% as male and 2.4% as other. Regarding ethnicity, 17.8% of participants were from minority backgrounds. Around one third of participants received free school meals. Overall, 31.0% of participants were from the most deprived IMD quintile in contrast to only 8.5% from the least deprived quintile. Broadly speaking, the intervention and control groups shared similar socio-demographic profiles except that there was a higher percentage of participants from minority ethnic backgrounds in the control group (21.9% vs 14.5%), and a higher percentage of participants from the most deprived IMD quintile in the intervention group (35.5% vs 25.4%).

### Primary outcomes

Figure 2a shows the predicted trajectories by intervention group from the growth curve model on total SDQ difficulties (see also supplementary material S6 for the table). For the intervention group, total SDQ difficulties decreased by 0.15 SD (95% CI: -0.25 to -0.04, p=0.006) between baseline and six-month follow-up. In contrast, there was little change by six-month follow up for the control group (slope: 0.01, 95% CI: -0.10 to 0.13, p=0.822). Examining specific SDQ domains (supplementary material S6 & S7), we found significant or marginally significant group differences in the rate of change in conduct problems, hyperactivity and peer relationship domains, but no difference in the emotional symptoms domain. Figure 2b shows the predicted trajectories of prosocial behaviour, which is captured within the SDQ but is not part of the total SDQ score (see also S6). The intervention and control groups showed significantly different trajectories between baseline and six-month (slope difference: 0.20, 95% CI: 0.02 to 0.38, p=0.032). Specifically, the control group experienced a decrease in prosocial behaviours (slope: -0.13, 95% CI: -0.26 to 0.01, p=0.066) in contrast to a slight increase (not statistically significant) for the intervention group (slope: 0.07, 95% CI: -0.05 to 0.19, p=0.242).

**Figure 2.**
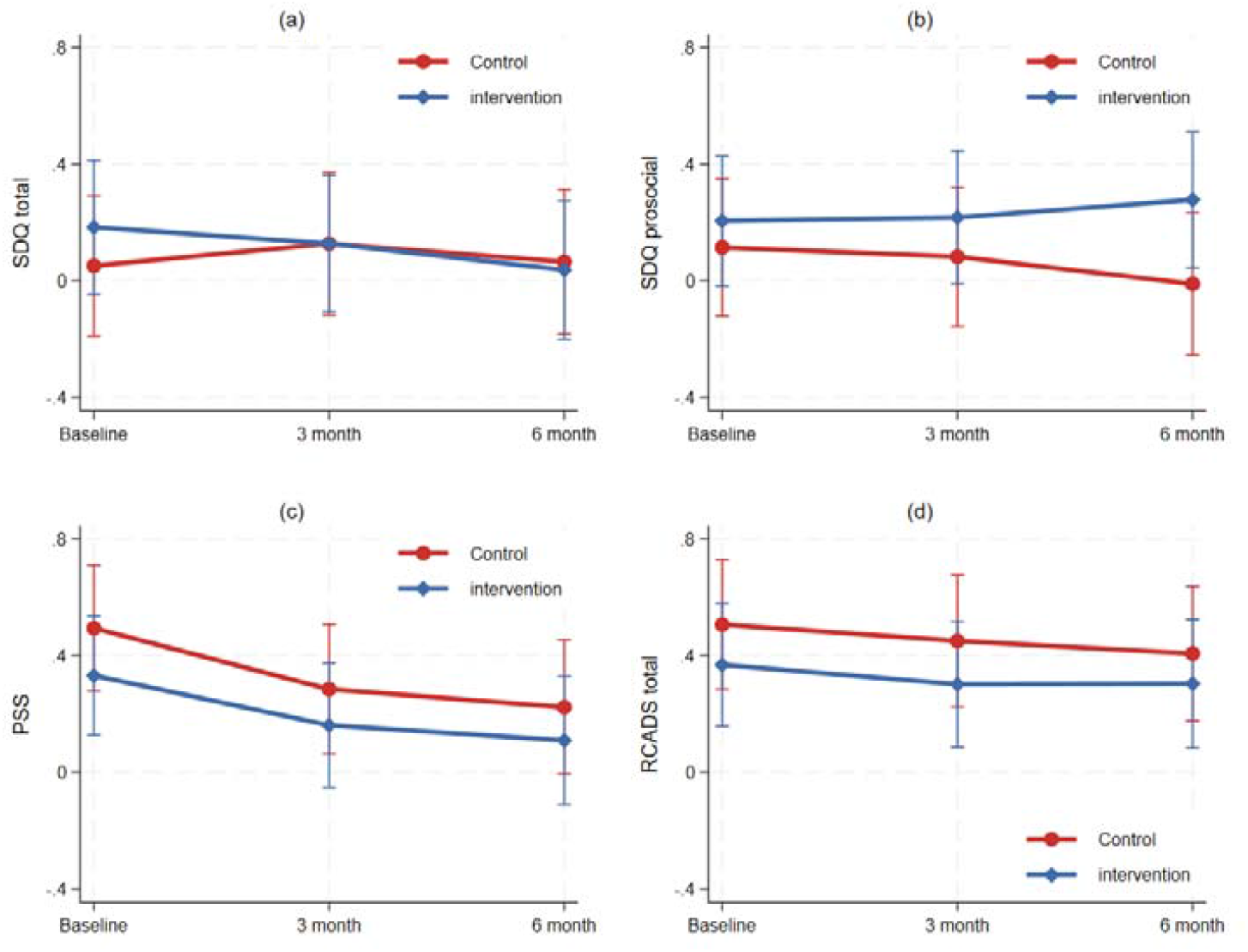
Estimated total SDQ difficulties, SDQ prosocial behaviour, stress (PSS) and mental health (RCADS) across different times points by intervention groups (reference characteristics: female, white ethnicity, no school meals, aged 15, third IMD quintile)

Figure 2c shows the predicted trajectories of perceived stress which decreased for both control and intervention group (see also S8). There was no evidence that the rate of change differed by intervention group (slope difference: 0.05, 95% CI: -0.17 to 0.27, p=0.664). Similarly, no group difference was found in anxiety and depression symptoms (slope difference: 0.03, 95% CI: -0.13 to 0.20, p=0.686; Table S8 & Figure S9).

### Secondary outcomes

Figure 3a shows the predicted trajectories of resilience by intervention group (see also S10). While there was little change between baseline and six-month for the control group (slope: 0.01, 95% CI: -0.11 to 0.13, p=0.868), resilience increased by 0.24 SD for the intervention group during the same follow-up period (95% CI: 0.13 to 0.35, p<0.001). Exploring specific domains of resilience, we found significant or marginally significant group differences in the rate of change in participation in community life, self-esteem and problem solving; whereas no difference was found in community connection, empathy or goals and aspirations (S10 & S11).

**Figure 3.**
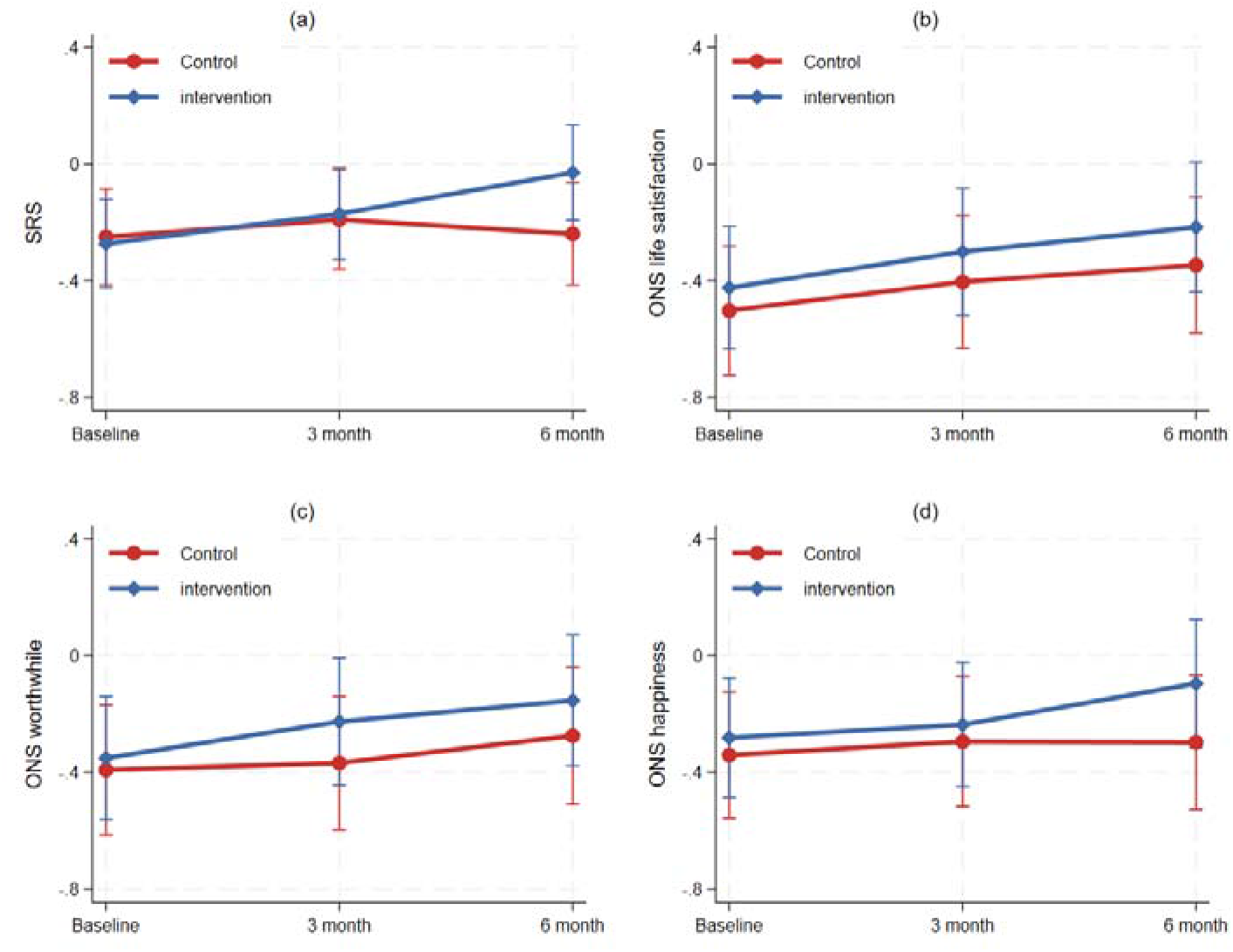
Estimated resilience (SRS) and ONS wellbeing measures across different times points by intervention groups (reference characteristics: female, white ethnicity, no school meals, aged 15, third IMD quintile)

Figures 3b-3d show the predicted trajectories of ONS wellbeing measures (see also S12). Life satisfaction increased between baseline and six-month follow up for both control and intervention groups, with little difference in the rate of change (slope difference: 0.05, 95% CI: -0.14 to 0.25, p=0.600). Sense of worthwhile also increased for both groups, specifically, by 0.12 SD for the control group (95% CI: -0.03 to 0.27, p=0.126) and by 0.20 SD for the intervention group (95% CI: 0.06 to 0.34, p=0.005). However, the group difference in the rate of change was not statistically significant (slope difference: 0.08, 95% CI: -0.12 to 0.28, p=0.442). For happiness, there was little change between baseline and 6-month for the control group (slope: 0.04, 95% CI: - 0.12 to 0.21, p=0.594). In contrast, happiness increased by 0.19 SD during the same follow-up period for the intervention group (95% CI: 0.04 to 0.33, p=0.014). However, it is of note that the group difference in the rate of change was not statistically significant (slope difference: 0.14, 95% CI: -0.08 to 0.36, p=0.203).

## Discussion

Youth SP delivered during CAMHS waiting periods appeared feasible and appropriate within routine CAMHS, with practitioners reporting high acceptability and feasibility, and young people reporting generally neutral to positive views of the intervention. Young people who received SP experienced improvements in psychosocial functioning and resilience. Relative to usual waitlist care, receipt of SP was associated with reductions in total difficulties, driven by improvements in conduct problems, hyperactivity/inattention, and peer problems, alongside increased prosocial behaviour. Resilience also improved across domains related to community participation, self-esteem, and problem-solving. In contrast, no clear between-group differences were observed for anxiety or depressive symptoms, or perceived stress. Taken together, these findings suggest that youth SP may exert short-term effects on day-to-day functioning and protective factors during waiting, rather than producing rapid symptom reduction for internalising difficulties.

This pattern is consistent with the broader youth SP evidence base, which indicates that reasons for referral in youth SP tend to focus around social and relational factors, as well as general mental health and wellbeing, with measures capturing constructs such as loneliness, connection, social skills and relationships [35]. Evidence reviews of youth SP, whilst still in their infancy, point towards a slightly more consistent evidence base for wellbeing outcomes [22, 23]. Whilst functional outcomes are more novel in this context, there is support for this within the adult literature [17, 18]. The present findings extend this evidence by demonstrating functional and resilience benefits within a CAMHS-referred cohort, a population characterised by clinically recognised need and heightened vulnerability to the destabilising effects of delayed access [7–10].

Qualitative findings from Wellbeing While Waiting provide further context for interpreting why improvements were observed in psychosocial functioning and resilience but not in internalising symptom measures. This work identified interlinked mechanisms centred on relational trust, youth autonomy and agency, behavioural activation, and facilitated social connection, with engagement shaped by contextual constraints such as complexity of needs and the accessibility of suitable community resources [26]. This mechanism profile is consistent with observed changes, in the present study, in behavioural difficulties and resilience outcomes, which capture behavioural regulation, peer functioning, participation, and coping resources. Moreover, the absence of short-term effects on symptom outcomes may be theoretically congruent with the intended function of SP. If SP primarily acts by strengthening social interactions, re-establishing routine and activity patterns, enhancing motivation, and increasing access to psychosocial resources, early impacts may plausibly be expected to emerge first in functioning and resilience domains rather than in rapid reductions in anxiety or depressive symptomatology, with downstream improvements in anxiety and depression potentially occurring over longer timeframes that future studies should investigate.

This interpretation is reinforced by broader theory on how community engagement and enjoyable activities influence mental health. The Multi-level Leisure Mechanisms Framework describes a range of behavioural, social, and psychological processes through which participation in activities may affect health, including behavioural activation, motivation and habit formation, social contact and bonding, and psychological processes such as coping, meaning in life, autonomy, and emotion regulation [36]. Importantly, the framework distinguishes between mechanisms that may be activated immediately and those that are likely to accrue over time, and emphasises the role of measurement choices and follow-up duration in capturing the mechanisms and outcomes most plausibly affected [36]. The waiting context is therefore likely to be central to interpreting impacts. Evidence on young people’s experiences of waiting for child and adolescent mental health services suggests that prolonged waits can constitute an active period of harm, characterised by uncertainty, strain, and risks of deterioration and disengagement, especially when support offered is incongruent with young people’s needs and preferences’ [7–10] Youth SP may function as an interim stabilising offer, supporting engagement with community-based protective resources, giving young people choice and control over their care, and providing relational continuity during a period of limited clinical input. Improvements in peer problems and prosocial behaviour are consistent with a buffering role against withdrawal and isolation during waiting, while resilience gains may reflect increased coping capacity and perceived competence in managing day-to-day challenges.

This study consisted of a large multi-site evaluation of youth SP in a policy relevant, high priority, population. Key strengths include the study’s sample size and multi-site delivery enhancing external validity, while the hybrid type II implementation–effectiveness design increases the translational value of the findings by situating outcome change within a real-world pathway. A further strength was the co-development of outcome measurement with a Youth Advisory Group, which helped ensure that core functional domains were captured using developmentally appropriate, youth-friendly measures. This strengthens content and face validity and acceptability, increasing the likelihood that observed changes reflect meaningful improvements in young people’s day-to-day functioning during the waiting period. Limitations primarily relate to internal validity and interpretability. The non-randomised design means residual confounding and secular changes cannot be ruled out, even though the temporal counterfactual within sites strengthens confidence in the observed between-group differences. Moreover, heterogeneity in delivery models across sites (e.g., embedded vs hosted LWs; remote-only delivery) and reliance on self-report outcomes may have diluted effects and limit inferences about which components and contextual conditions drive impact; however, the pragmatic design intentionally reflects real-world variation in how SP services are delivered.

Clinically, these findings support youth SP as a credible adjunctive bridging offer during CAMHS waiting periods, particularly for young people whose presentations centre on impaired functioning, social withdrawal, reduced participation, behavioural dysregulation, and weakened coping resources. Improvements across SDQ domains and resilience outcomes suggest that SP may contribute to stabilisation and strengthening of protective factors during a high-risk period of delayed access. The findings are an important response to the growing literature on waiting-list interventions for children and young people using CAMHS, which highlights both the clinical importance of the waiting period and the need for feasible interim supports while specialist care is pending [11]. However, the absence of short-term reductions in anxiety and depressive symptoms indicates that youth SP should not be positioned as a substitute for evidence-based psychological treatments, particularly around internalising disorders; a view which is echoed within best practice guidelines for youth SP [37].

In terms of clinical and policy implications, the results strengthen the case for youth SP as part of a system response to waiting-list harms, but underline that impact is inseparable from implementation quality and equity of access. Best practice guidance further indicates that effective youth SP requires sustained workforce capacity, supervision and safeguarding pathways, plus practical support to overcome barriers such as transport, cost and uneven local provision; scaling referral models without addressing these conditions risks undermining relational quality and widening inequalities [37]. Overall, interpreted alongside mechanistic, best-practice and implementation evidence, the findings support positioning youth SP as a relational, strengths-based intervention that can stabilise young people and strengthen protective resources during waiting, while reinforcing the need for equitable delivery models and integration with timely access to specialist mental health care.

## Acknowledgements

This paper is independent research supported by the National Institute for Health Research ARC North Thames. The views expressed in this publication are those of the author(s) and not necessarily those of the National Institute for Health Research or the Department of Health and Social Care.

## Declarations

### Ethics

Ethics approval for the trial was granted by the NHS Research Ethics Committee (Ref 22/WS/0184).

### Consent

Young people aged 16–18 provided informed consent. Young people aged 11–15 gave assent, with caregivers providing informed consent for both their own and their child’s participation.

### Data availability

The datasets generated and/or analysed during the current study are not publicly available due to governance restrictions but are available from the corresponding author on reasonable request and subject to appropriate approvals.

### Author contributions

DF, DH, JD, ABu and FB conceived and designed the study. DH, JW, ABu, LS, HS, JP, ABr, and EH contributed to the acquisition and curation of data. FBu conducted the analysis with input from all other authors. DH and DF drafted the manuscript. All authors reviewed, revised, and approved of the final version for submission.

## References

1. Liu Y, Ren Y, Liu C, et al (2025) Global burden of mental disorders in children and adolescents before and during the COVID-19 pandemic: evidence from the Global Burden of Disease Study 2021. Psychol Med 55:e90. 10.1017/S0033291725000649

2. NHS Digital (2023) Mental Health of Children and Young People in England, 2023 - wave 4 follow up to the 2017 survey. In: NHS Digital. https://digital.nhs.uk/data-and-information/publications/statistical/mental-health-of-children-and-young-people-in-england/2023-wave-4-follow-up#. Accessed 16 Feb 2024

3. NHS England (2025) Children and young people accessing mental health services. In: NHS England. https://app.powerbi.com/view?r=eyJrIjoiOTdjYzFiYTUtZmEwMi00ZTA2LTkxOGUtMDZmMmZjMThiZGNhIiwidCI6IjM3YzM1NGIyLTg1YjAtNDdmNS1iMjIyLTA3YjQ4ZDc3NGVlMyJ9. Accessed 18 Jan 2026

4. NHS Digital (2024) Mental Health Services Monthly Statistics, Performance

5. Department of Health and Department of Education (2017) Transforming Children and Young People’s Mental Health Provision: A Green Paper. London

6. Holt-White E, Lathamm K., Anders J, et al (2023) Wave 2 Initial Findings - Briefing No. 1. Mental and physical health. London

7. Han E, Burton A, Bradbury A, et al (2026) Experiences of youth and caregivers waiting for mental health services in the UK: a qualitative study to inform policy and practice. Eur Child Adolesc Psychiatry. 10.1007/s00787-025-02952-x

8. Subotic-Kerry M, Borchard T, Parker B, et al (2025) While they wait: a cross-sectional survey on wait times for mental health treatment for anxiety and depression for adolescents in Australia. BMJ Open 15:e087342. 10.1136/bmjopen-2024-087342

9. Punton G, Dodd AL, McNeill A (2022) ‘You’re on the waiting list’: An interpretive phenomenological analysis of young adults’ experiences of waiting lists within mental health services in the UK. PLoS One 17:e0265542. 10.1371/journal.pone.0265542

10. Biringer E, Sundfør B, Davidson L, et al (2015) Life on a waiting list: How do people experience and cope with delayed access to a community mental health center? Scandinavian Psychologist 2:. 10.15714/scandpsychol.2.e6

11. Valentine AZ, Hall SS, Sayal K, Hall CL (2024) Waiting-list interventions for children and young people using child and adolescent mental health services: a systematic review. BMJ Mental Health 27:e300844. 10.1136/bmjment-2023-300844

12. National Academy for Social Prescribing (2021) What is Social Prescribing? In: National Academy for Social Prescribing,. https://socialprescribingacademy.org.uk/what-is-social-prescribing/#:~:text=Social%20prescribing%20connects%20people%20to,on%20what%20works%20for%20them. https://www.england.nhs.uk/personalisedcare/workforce-and-training/social-prescribing-link-workers/#:~:text=Social%20prescribing%20link%20workers%20connect,housing%2C%20financial%20and%20welfare%20advice. Accessed 13 Aug 2023

13. NHS England (2021) Social Prescribing Link Workers. In: NHS England,. https://www.england.nhs.uk/personalisedcare/workforce-and-training/social-prescribing-link-workers/#:~:text=Social%20prescribing%20link%20workers%20connect,housing%2C%20financial%20and%20welfare%20advice. Accessed 13 Aug 2023

14. NHS (2019) The NHS Long Term Plan. London

15. Dayson C, Bashir N (2014) The social and economic impact of the Rotherham Social Prescribing Pilot: main evaluation report. . Sheffield

16. Kimberlee R, Bertotti M, Dayson C, et al (2022) The economic impact of social prescribing. London

17. Chatterjee HJ, Camic PM, Lockyer B, Thomson LJ (2018) Non-clinical community interventions: a systematised review of social prescribing schemes. Arts & Health 10:97–123

18. Bickerdike L, Booth A, Wilson PM, et al (2017) Social prescribing: less rhetoric and more reality. A systematic review of the evidence. BMJ Open e013384

19. Aggar C, Thomas T, Gordon C, et al (2021) Social Prescribing for Individuals Living with Mental Illness in an Australian Community Setting: A Pilot Study. Community Ment Health J 57:189–195. 10.1007/s10597-020-00631-6

20. Dayson C, Painter J, Bennett E (2020) Social prescribing for patients of secondary mental health services: emotional, psychological and social well-being outcomes. J Public Ment Health 19:271–279. 10.1108/JPMH-10-2019-0088

21. Bertotti M, Hayes D, Berry V, et al (2022) Social prescribing for children and young people. Lancet Child Adolesc Health 6:835–837. 10.1016/S2352-4642(22)00248-6

22. Hayes D, Jarvis-Beesley P, Mitchell D, et al (2023) The impact of social prescribing on children and young people’s mental health and wellbeing’. . London

23. Muhl C, Cornish E, Zhou XA, et al (2025) Social Prescribing for Children and Youth: A Scoping Review. Health Soc Care Community 2025:. 10.1155/hsc/5265529

24. NHS England (2020) Social Prescribing . In: NHS England,. https://www.england.nhs.uk/personalisedcare/social-prescribing/#:~:text=Social%20prescribing%20is%20an%20all,who%20are%20lonely%20or%20isolated. Accessed 13 Aug 2023

25. Fancourt D, Burton A, Bu F, et al (2023) Wellbeing while waiting evaluating social prescribing in CAMHS: study protocol for a hybrid type II implementation-effectiveness study. BMC Psychiatry 23:328. 10.1186/s12888-023-04758-0

26. Han E, Bradbury A, Burton A, et al (2026) Understanding how and why social prescribing supports young people waiting for mental health services: a qualitative study of mechanisms and contextual factors

27. Schulz KF, Altman DG, Moher D (2010) CONSORT 2010 Statement: updated guidelines for reporting parallel group randomised trials. BMJ 340:c332–c332. 10.1136/bmj.c332

28. Zwarenstein M, Treweek S, Gagnier JJ, et al (2008) Improving the reporting of pragmatic trials: an extension of the CONSORT statement. BMJ 337:a2390–a2390. 10.1136/bmj.a2390

29. Goodman R (1997) The strengths and difficulties questionnaire: A research note. Journal of Child Psychology and Psychiatry 38:581–586

30. Radez J, Waite P, Chorpita B, et al (2021) Using the 11-item Version of the RCADS to Identify Anxiety and Depressive Disorders in Adolescents. Res Child Adolesc Psychopathol 49:1241–1257. 10.1007/s10802-021-00817-w

31. Demkowicz O, Panayiotou M, Ashworth E, et al (2020) The Factor Structure of the 4-Item Perceived Stress Scale in English Adolescents. European Journal of Psychological Assessment 36:913–917. 10.1027/1015-5759/a000562

32. Sun J, Stewart D (2007) Development of population-based resilience measures in the primary school setting. Health Educ Res 7:575–599

33. Jordan A, Rees E (2020) Children’s well-being indicator review, UK: 2020. In: Office for National Statistics . https://www.ons.gov.uk/peoplepopulationandcommunity/wellbeing/articles/childrenswellbeingindicatorreviewuk2020/2020-09-02. Accessed 18 Jan 2023

34. Weiner, B. J., Lewis, C. C., Stanick, C., Powell, B. J., Dorsey, C. N., Clary, A. S., … & Halko H (2017) Psychometric assessment of three newly developed implementation outcome measures. Implementation Science 12:108

35. Mitchell SB, Cartwright L, Gude A, et al (2025) The use of social prescribing and community-based wellbeing activities as a potential prevention and early intervention pathway to improve adolescent emotional and social development: a systematic mapping review. BMC Public Health 25:3495. 10.1186/s12889-025-24413-5

36. Fancourt D, Aughterson H, Finn S, et al (2021) How leisure activities affect health: a narrative review and multi-level theoretical framework of mechanisms of action. Lancet Psychiatry 8:329–339. 10.1016/S2215-0366(20)30384-9

37. Hayes D, Arslanovski N, Fancourt D, et al (2026) Best Practice Guidelines: Child and Youth Social Prescribing. London

