## Supplementary Materials for "Wellbeing While Waiting: Effectiveness and implementation of youth social prescribing for young people awaiting CAMHS support"

Supplementary material

S1: Site level implementation dates

S2: Details of study design choice.

S3: CONSORT CHECKLISTS

S4: Site level social prescribing model

S5: Figure SF1a-d: Acceptability, appropriateness and feasibility scores from participants

S6: Table ST1: Results from growth curve models on SDQ total score and subdomains

S7: Figure SF2. Estimated SDQ subdomains across different time points by intervention group

S8: Table ST2: Results from growth curve models on PSS, RCADS total score and subdomains

S9: Figure SF3. Estimated RCADS subdomains across different time points by intervention groups

S10: Table ST3 Results from growth curve models on SRS total score and subdomains

S11: Figure SF4. Estimated SRS subdomains across different time points by intervention groups

S12: Table ST4 Results from growth curve models on ONS wellbeing measures

**S1: Site level implementation dates**

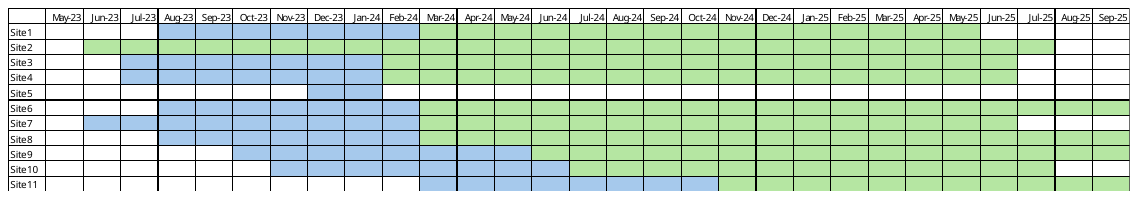

**S2: Details of study design choice.**

This study followed a pragmatic non-randomised stepped wedge design. This design was chosen over a traditional individual or cluster randomised controlled trial for several reasons, including 1) to avoid unethical provision of a potentially beneficial service to some but not all people currently on a CAMHS waiting list, 2) to increase uptake of a novel complex intervention where cluster randomisation could lead to study non-participation, 3) to allow distribution of resources across multiple sites when rolling out a complex intervention through phased implementation delivery, 4) to effectively capture the real-world impact of a complex intervention (i.e. one with multiple component parts), and 5) to allow evaluations of the implementation of a complex intervention alongside its efficacy. Stepped wedge approaches blend constraints relating to complex intervention implementation with attempts to provide rigorous scientific evaluations ^1–4^.

Whereas traditional stepped wedge designs involve randomly allocating sites to crossover from control to intervention, our study was a non-randomised stepped-wedge design. This non-randomised design has been used in interventions before, especially those that depend on roll-outs happening at specific timepoints where randomisation is not feasible^5^, or those where there remain ethical concerns about randomising even at a cluster level. It is also pragmatic in studies involving the delivery of new complex pathways or studies simultaneously looking at implementation as the crossover of sites at different times gives an opportunity for learning of how to improve delivery in future sites^6^. Further, it can help to address issues surrounding how to stick to specific crossover dates when dealing with multiple busy sites^6^. Thus, non-randomised stepped wedge trials can provide important quasi-experimental designs that provide nuanced approaches to the simultaneous implementation and evaluation of interventions.

Instead of randomisation, our study involved a pragmatic approach driven by recruitment targets to determine when sites crossed over to offering the social prescribing intervention. Once sites opened, they recruited the control allocation first. To further minimise ethical effects of delaying a potentially beneficial activity^6^, during this control period, sites finalised their social prescribing pathway ready for roll out. This design had the pragmatic effect of removing implementation lag as each site was ready to crossover as soon as they had hit their control recruitment target, and rendering the study design ‘complete’ rather than ‘incomplete’^7^. Once sites had hit their control target, providing their social prescribing pathway was ready to go, they crossed over to delivering social prescribing. As sites all entered the study at different timepoints and had different recruitment targets, this created natural variability in the step length, resulting in the modified step-wedge design shown below, with informal ‘clusters’ who crossed over at similar timepoints.

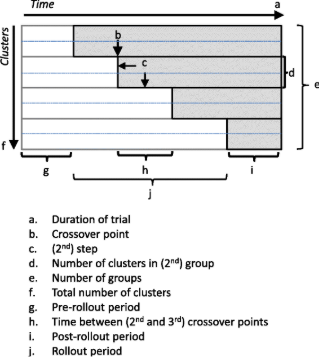

https://trialsjournal.biomedcentral.com/articles/10.1186/s13063-015-0842-7

We anticipated 8-10 sites in our study, although recruited 11 sites to mitigate against site drop-out. The design of the stepped wedge was ‘continuous recruitment with short exposure’^7^: individuals became eligible continuously over the course of the study and participated for their 6-month study period before then leaving the study. As individuals had to be recruited within the first four weeks of being triaged within CAMHS, they could only take part in one or other condition (i.e. control or social prescribing). For the small number of individuals who were recruited while their site was under control conditions but who were still within the four weeks when their site crossed over to offering social prescribing, the condition they were recruited under was the condition they continued to receive. Once individuals had participated in the trial, they were ineligible to participate again (even if they received a separate referral to CAMHS which resulted in another triage). This design had the added advantage of precluding any carry-over effects. Link workers did not work with individuals unless the site had crossed over (avoiding any bias in their engagement with the young people as a result of training in anticipation of the social prescribing being offered) and individuals had not been on a waiting list long enough to experience the specific negative psychological ‘waiting list effects’ that the trial was attempting to reduce.

The full description of the design is T4/D2/M2. This means that the timing of the exposure (T) was such that no individual was exposed at the start but they were first exposed in a continuous and gradual process, as they were recruited in and met with a link worker (if in the intervention arm). The duration (D) was of varying length for each individual depending on the social prescribing intervention that was chosen with them in consultation with the link worker. However, the measurement (M) was consistent across individuals - repeated measurement at times linked to the start of their exposure. We chose separate steps for each study site to meet the pragmatic design of the trial. However, this also has the benefit of resulting in greater statistical power^8^.

Stepped wedge designs can introduce bias by design due to temporal trends outside of the study. However, we took some steps to mitigate against this. First, in line with some previous stepped wedge studies, one site was recruited that already had a social prescribing pathway active, so did not collect any control data^9^. This had the benefit of providing simultaneous data on social prescribing at the same time that all the other sites were collecting control data, allowing an assessment of whether external biases affected the controls. Second, we had a large number of steps (one per site), which can help to model trends occurring outside the study over time^10^. Third, we collected a broad range of socio-demographic information of young people and their families, allowing us to inspect and control for systematic differences between control and intervention groups. Consequently, secular trends can be adjusted for and estimated in the analysis.

References

1.	Stepped-Wedge Trials | Quantifying impact | LSHTM. https://www.lshtm.ac.uk/research/centres/centre-evaluation/stepped-wedge-trials.

2.	Hemming, K., Haines, T. P., Chilton, P. J., Girling, A. J. & Lilford, R. J. The stepped wedge cluster randomised trial: rationale, design, analysis, and reporting. BMJ 350, h391 (2015).

3.	Greenhalgh, T. & Papoutsi, C. Studying complexity in health services research: desperately seeking an overdue paradigm shift. BMC Med. 16, 95 (2018).

4.	Marchal, B. et al. Realist RCTs of complex interventions – An oxymoron. Soc. Sci. Med. 94, 124–128 (2013).

5.	Highfield, L. et al. A non-randomized controlled stepped wedge trial to evaluate the effectiveness of a multi-level mammography intervention in improving appointment adherence in underserved women. Implement. Sci. 10, 143 (2015).

6.	Prost, A. et al. Logistic, ethical, and political dimensions of stepped wedge trials: critical review and case studies. Trials 16, 351 (2015).

7.	Copas, A. J. et al. Designing a stepped wedge trial: three main designs, carry-over effects and randomisation approaches. Trials 16, 352 (2015).

8.	Baio, G. et al. Sample size calculation for a stepped wedge trial. Trials 16, 354 (2015).

9.	Fuller, C. et al. The Feedback Intervention Trial (FIT) — Improving Hand-Hygiene Compliance in UK Healthcare Workers: A Stepped Wedge Cluster Randomised Controlled Trial. PLOS ONE 7, e41617 (2012).

10.	Hemming, K. & Taljaard, M. Reflection on modern methods: when is a stepped-wedge cluster randomized trial a good study design choice? Int. J. Epidemiol. 49, 1043–1052 (2020)

**S3: CONSORT CHECKLISTS**

**A: CONSORT 2025 checklist of information to include when reporting a randomised trial***

| **Section / Topic** | **No** | **CONSORT 2025 checklist item description** | **Reported on page no.** |
| --- | --- | --- | --- |
| **Title and abstract** | | |  |
| Title and structured abstract | 1a | Identification as a randomised trial | NA - Note: describe as non-randomised controlled trial |
|  | 1b | Structured summary of the trial design, methods, results, and conclusions | 2 |
| **Open science** | | |  |
| Trial registration | 2 | Name of trial registry, identifying number (with URL) and date of registration | 6 |
| Protocol and statistical analysis plan | 3 | Where the trial protocol and statistical analysis plan can be accessed | 6 |
| Data sharing | 4 | Where and how the individual de-identified participant data (including data dictionary), statistical code and any other materials can be accessed | 27 |
| Funding and conflicts of interest | 5a | Sources of funding and other support (e.g., supply of drugs), and role of funders in the design, conduct, analysis and reporting of the trial | 26 |
|  | 5b | Financial and other conflicts of interest of the manuscript authors | 26 |
| **Introduction** | | |  |
| Background and rationale | 6 | Scientific background and rationale | 3-5 |
| Objectives | 7 | Specific objectives related to benefits and harms | 5 |
| **Methods** | | |  |
| Patient and public involvement | 8 | Details of patient or public involvement in the design, conduct and reporting of the trial | 6 |
| Trial design | 9 | Description of trial design including type of trial (e.g., parallel group, crossover), allocation ratio, and framework (e.g., superiority, equivalence, non-inferiority, exploratory) | 6-7 |
| Changes to trial protocol | 10 | Important changes to the trial after it commenced including any outcomes or analyses that were not prespecified, with reason | 10 |
| Trial setting | 11 | Settings (e.g., community, hospital) and locations (e.g., countries, sites) where the trial was conducted | 7 |
| Eligibility criteria | 12a | Eligibility criteria for participants | 8 |
|  | 12b | If applicable, eligibility criteria for sites and for individuals delivering the interventions (e.g., surgeons, physiotherapists) | 7-9 |
| Intervention and comparator | 13 | Intervention and comparator with sufficient details to allow replication. If relevant, where additional materials describing the intervention and comparator (e.g., intervention manual) can be accessed | 8-9 |
| Outcomes | 14 | Pre-specified primary and secondary outcomes, including the specific measurement variable (e.g., systolic blood pressure), analysis metric (e.g., change from baseline, final value, time to event), method of aggregation (e.g., median, proportion), and time point for each outcome | 9-10 |
| Harms | 15 | How harms were defined and assessed (e.g., systematically, non-systematically) | 13 |
| Sample size | 16a | How sample size was determined, including all assumptions supporting the sample size calculation | S2 |
|  | 16b | Explanation of any interim analyses and stopping guidelines | n/a |
| Randomisation: |  |  |  |
| Sequence generation | 17a | Who generated the random allocation sequence and the method used | n/a |
|  | 17b | Type of randomisation and details of any restriction (e.g., stratification, blocking and block size) | n/a |
| Allocation concealment mechanism | 18 | Mechanism used to implement the random allocation sequence (e.g., central computer/telephone; sequentially numbered, opaque, sealed containers), describing any steps to conceal the sequence until interventions were assigned | n/a |
| Implementation | 19 | Whether the personnel who enrolled and those who assigned participants to the interventions had access to the random allocation sequence | n/a |
| Blinding | 20a | Who was blinded after assignment to interventions (e.g., participants, care providers, outcome assessors, data analysts) | n/a |
|  | 20b | If blinded, how blinding was achieved and description of the similarity of interventions | n/a |
| Statistical methods | 21a | Statistical methods used to compare groups for primary and secondary outcomes, including harms | 10-11 |
|  | 21b | Definition of who is included in each analysis (e.g., all randomised participants), and in which group | 14 |
|  | 21c | How missing data were handled in the analysis | 10 |
|  | 21d | Methods for any additional analyses (e.g., subgroup and sensitivity analyses), distinguishing prespecified from post-hoc | 17-19 |
| **Results** | | |  |
| Participant flow, including flow diagram | 22a | For each group, the numbers of participants who were randomly assigned, received intended intervention, and were analysed for the primary outcome | Figure 1 |
|  | 22b | For each group, losses and exclusions after randomisation, together with reasons | 14 |
| Recruitment | 23a | Dates defining the periods of recruitment and follow-up for outcomes of benefits and harms | 12 |
|  | 23b | If relevant, why the trial ended or was stopped | n/a |
| Intervention and comparator delivery | 24a | Intervention and comparator as they were actually administered (e.g., where appropriate, who delivered the intervention/comparator, how participants adhered, whether they were delivered as intended [fidelity]) | 12-13 |
|  | 24b | Concomitant care received during the trial for each group | 9 |
| Baseline data | 25 | A table showing baseline demographic and clinical characteristics for each group | Table 1 |
| Numbers analysed,  outcomes and estimation | 26 | For each primary and secondary outcome, by group:   - the number of participants included in the analysis - the number of participants with available data at the outcome time point - result for each group, and the estimated effect size and its precision (such as 95% confidence interval) - for binary outcomes, presentation of both absolute and relative effect size | 15-19 |
| Harms | 27 | All harms or unintended events in each group | 13 |
| Ancillary analyses | 28 | Any other analyses performed, including subgroup and sensitivity analyses, distinguishing pre-specified from post-hoc | S6-S12 |
| **Discussion** | | |  |
| Interpretation | 29 | Interpretation consistent with results, balancing benefits and harms, and considering other relevant evidence | 19-22 |
| Limitations | 30 | Trial limitations, addressing sources of potential bias, imprecision, generalisability, and, if relevant, multiplicity of analyses | 23 |

*We strongly recommend reading this statement in conjunction with the CONSORT 2025 Explanation and Elaboration and/or the CONSORT 2025 Expanded Checklist for important clarifications on all the items. We also recommend reading relevant CONSORT extensions. See [www.consort-spirit.org](http://www.consort-spirit.org).

Citation: Hopewell S, Chan AW, Collins GS, Hróbjartsson A, Moher D, Schulz KF, et al. CONSORT 2025 Statement: updated guideline for reporting randomised trials. BMJ. 2025; 388:e081123. <https://dx.doi.org/10.1136/bmj-2024-081123>.

© 2025 Hopewell et al. This is an Open Access article distributed under the terms of the Creative Commons Attribution License (<https://creativecommons.org/licenses/by/4.0/>), which permits unrestricted use, distribution, and reproduction in any medium, provided the original work is properly cited.

**B: Checklist of items for reporting pragmatic trials**

| Section | Item | Standard CONSORT description | Extension for pragmatic trials | Page |
| --- | --- | --- | --- | --- |
| Title and abstract | 1 | How participants were allocated to interventions (eg, “random allocation,” “randomised,” or “randomly assigned”) |  |  |
| **Introduction** |  |  |  |  |
| Background | 2 | Scientific background and explanation of rationale | Describe the health or health service problem that the intervention is intended to address and other interventions that may commonly be aimed at this problem | 3-5 |
| **Methods** |  |  |  |  |
| Participants | 3 | Eligibility criteria for participants; settings and locations where the data were collected | Eligibility criteria should be explicitly framed to show the degree to which they include typical participants and/or, where applicable, typical providers (eg, nurses), institutions (eg, hospitals), communities (or localities eg, towns) and settings of care (eg, different healthcare financing systems) | 7-8 |
| Interventions | 4 | Precise details of the interventions intended for each group and how and when they were actually administered | Describe extra resources added to (or resources removed from) usual settings in order to implement intervention. Indicate if efforts were made to standardise the intervention or if the intervention and its delivery were allowed to vary between participants, practitioners, or study sites | 7-9 |
|  |  |  | Describe the comparator in similar detail to the intervention | 9 |
| Objectives | 5 | Specific objectives and hypotheses |  |  |
| Outcomes | 6 | Clearly defined primary and secondary outcome measures and, when applicable, any methods used to enhance the quality of measurements (eg, multiple observations, training of assessors) | Explain why the chosen outcomes and, when relevant, the length of follow-up are considered important to those who will use the results of the trial | 9-10 |
| Sample size | 7 | How sample size was determined; explanation of any interim analyses and stopping rules when applicable | If calculated using the smallest difference considered important by the target decision maker audience (the minimally important difference) then report where this difference was obtained | S2 |
| Randomisation—sequence generation | 8 | Method used to generate the random allocation sequence, including details of any restriction (eg, blocking, stratification) |  |  |
| Randomisation—allocation concealment | 9 | Method used to implement the random allocation sequence (eg, numbered containers or central telephone), clarifying whether the sequence was concealed until interventions were assigned |  |  |
| Randomisation—implementation | 10 | Who generated the allocation sequence, who enrolled participants, and who assigned participants to their groups |  |  |
| Blinding (masking) | 11 | Whether participants, those administering the interventions, and those assessing the outcomes were blinded to group assignment | If blinding was not done, or was not possible, explain why | N/A |
| Statistical methods | 12 | Statistical methods used to compare groups for primary outcomes; methods for additional analyses, such as subgroup analyses and adjusted analyses |  |  |
| **Results** |  |  |  |  |
| Participant flow | 13 | Flow of participants through each stage (a diagram is strongly recommended)—specifically, for each group, report the numbers of participants randomly assigned, receiving intended treatment, completing the study protocol, and analysed for the primary outcome; describe deviations from planned study protocol, together with reasons | The number of participants or units approached to take part in the trial, the number which were eligible, and reasons for non-participation should be reported | 12 and Figure 1 |
| Recruitment | 14 | Dates defining the periods of recruitment and follow-up |  |  |
| Baseline data | 15 | Baseline demographic and clinical characteristics of each group |  |  |
| Numbers analysed | 16 | Number of participants (denominator) in each group included in each analysis and whether analysis was by “intention-to-treat”; state the results in absolute numbers when feasible (eg, 10/20, not 50%) |  |  |
| Outcomes and estimation | 17 | For each primary and secondary outcome, a summary of results for each group and the estimated effect size and its precision (eg, 95% CI) |  |  |
| Ancillary analyses | 18 | Address multiplicity by reporting any other analyses performed, including subgroup analyses and adjusted analyses, indicating which are prespecified and which are exploratory |  |  |
| Adverse events | 19 | All important adverse events or side effects in each intervention group |  |  |
| **Discussion** |  |  |  |  |
| Interpretation | 20 | Interpretation of the results, taking into account study hypotheses, sources of potential bias or imprecision, and the dangers associated with multiplicity of analyses and outcomes |  |  |
| Generalisability | 21 | Generalisability (external validity) of the trial findings | Describe key aspects of the setting which determined the trial results. Discuss possible differences in other settings where clinical traditions, health service organisation, staffing, or resources may vary from those of the trial | 23 |
| Overall evidence | 22 | General interpretation of the results in the context of current evidence |  |  |

***Cite as:*** *Zwarenstein M, Treweek S, Gagnier JJ, Altman DG, Tunis S, Haynes B, Oxman AD, Moher D for the CONSORT and Pragmatic Trials in Healthcare (Practihc) group. Improving the reporting of pragmatic trials: an extension of the CONSORT statement. BMJ 2008; 337;a2390.*

**S4: Site level social prescribing model**

| **Site** | **Social Prescribing Model** | **Delivery mode** |
| --- | --- | --- |
| 1 | Hosted externally with formal CAMHS pathway integration | Mixed/in person available |
| 2 | Link Worker embedded in CAMHS | Mixed/in person available |
| 3 | Hosted externally with formal CAMHS pathway integration | Remote only |
| 4 | Hosted externally with formal CAMHS pathway integration | Mixed/in person available |
| 5 | Service did not commence social prescribing | N/A |
| 6 | Hosted externally with formal CAMHS pathway integration | Mixed/in person available |
| 7 | Hosted externally with formal CAMHS pathway integration | Mixed/in person available |
| 8 | Hosted externally with formal CAMHS pathway integration | Remote only |
| 9 | Hosted externally with formal CAMHS pathway integration | Mixed/in person available |
| 10 | Hosted externally with formal CAMHS pathway integration | Mixed/in person available |
| 11 | Link Worker embedded in CAMHS | Mixed/in person available |

**S5: Figure SF1a-d: Acceptability, appropriateness and feasibility scores from participants**

**51a: Youth AIM scores**

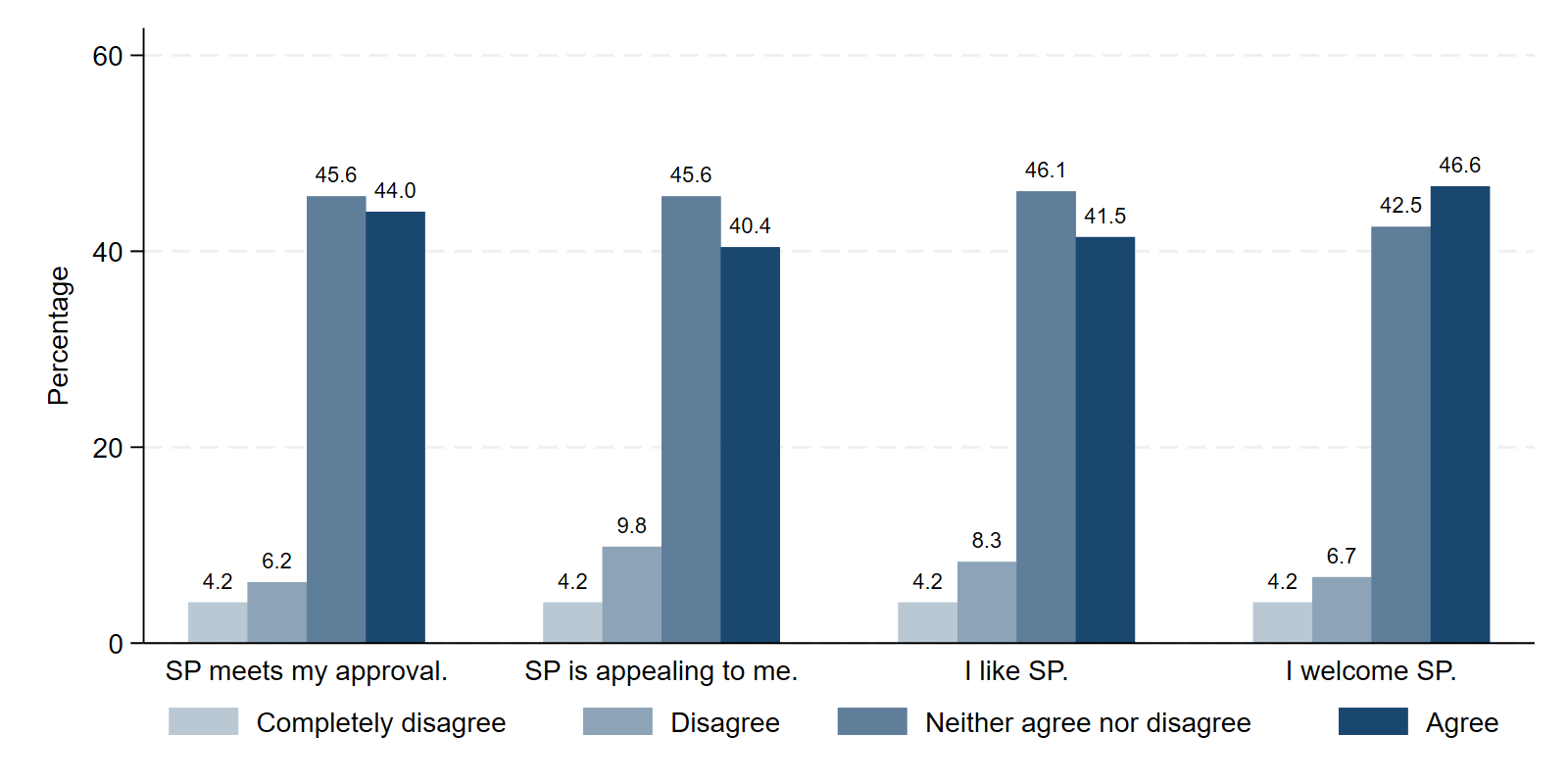

**51b: Staff and LW AIM scores**

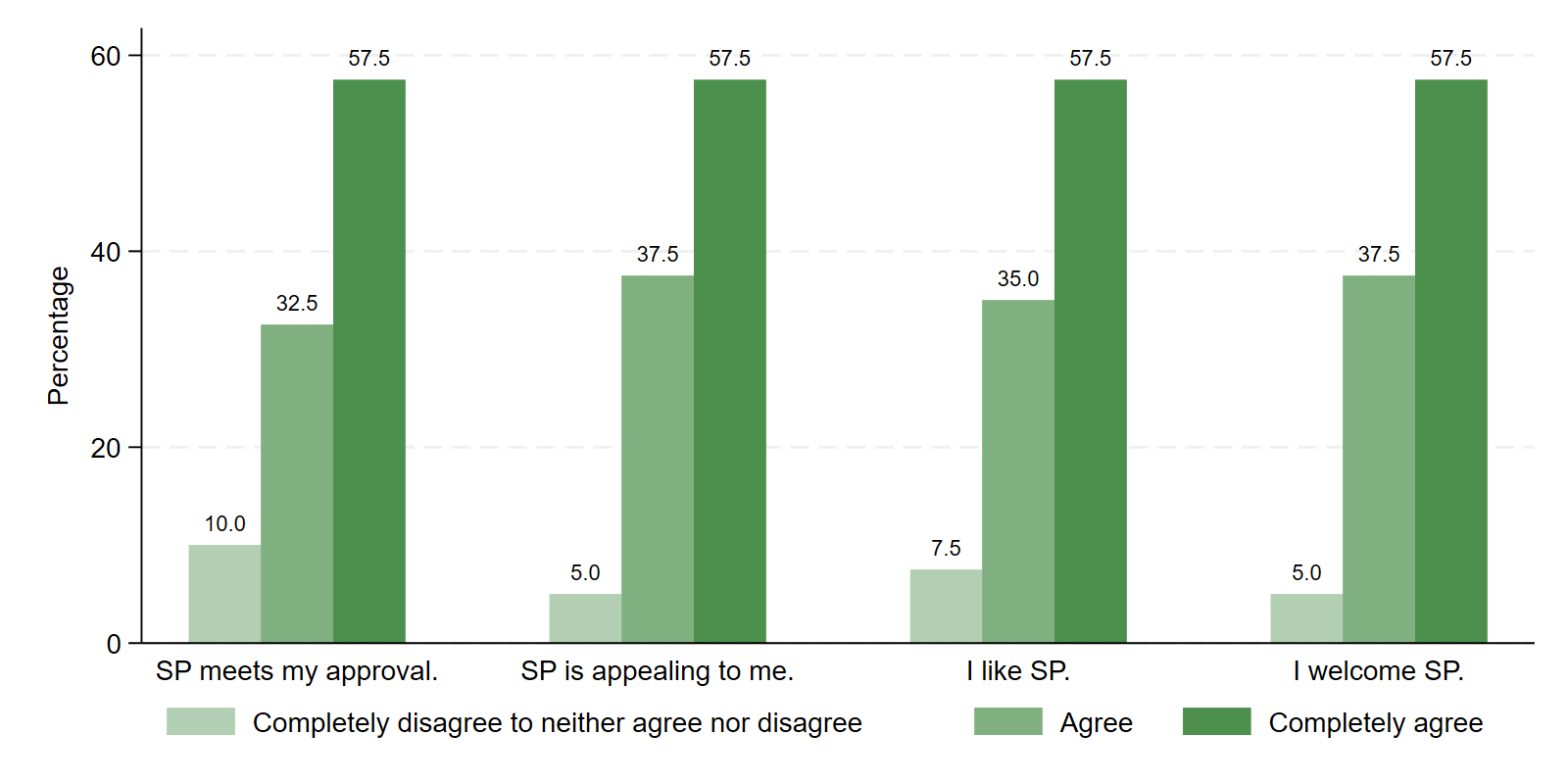

**51c: Staff and LW FIM scores**

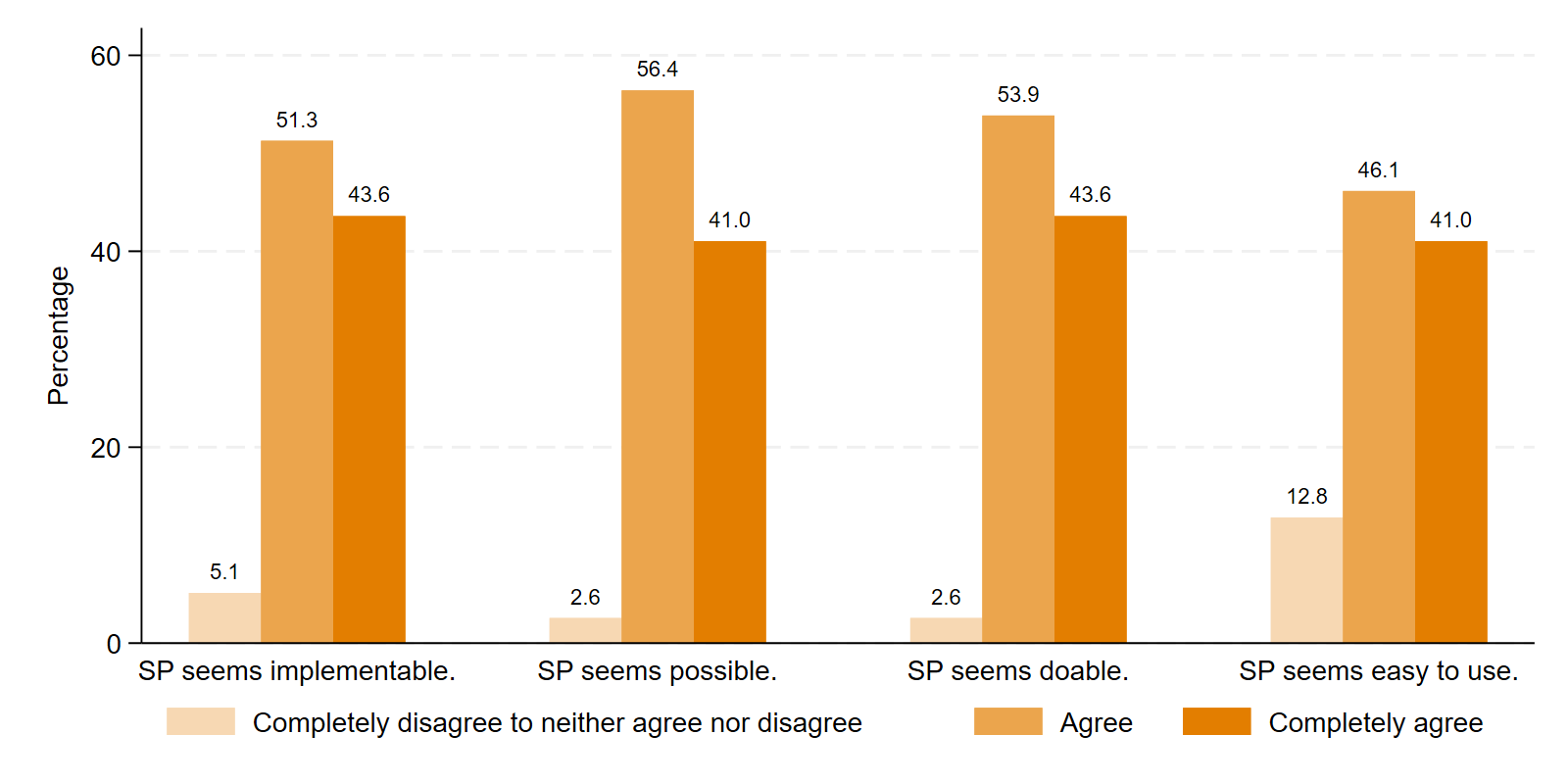

**51d: Staff and LW IAM scores**

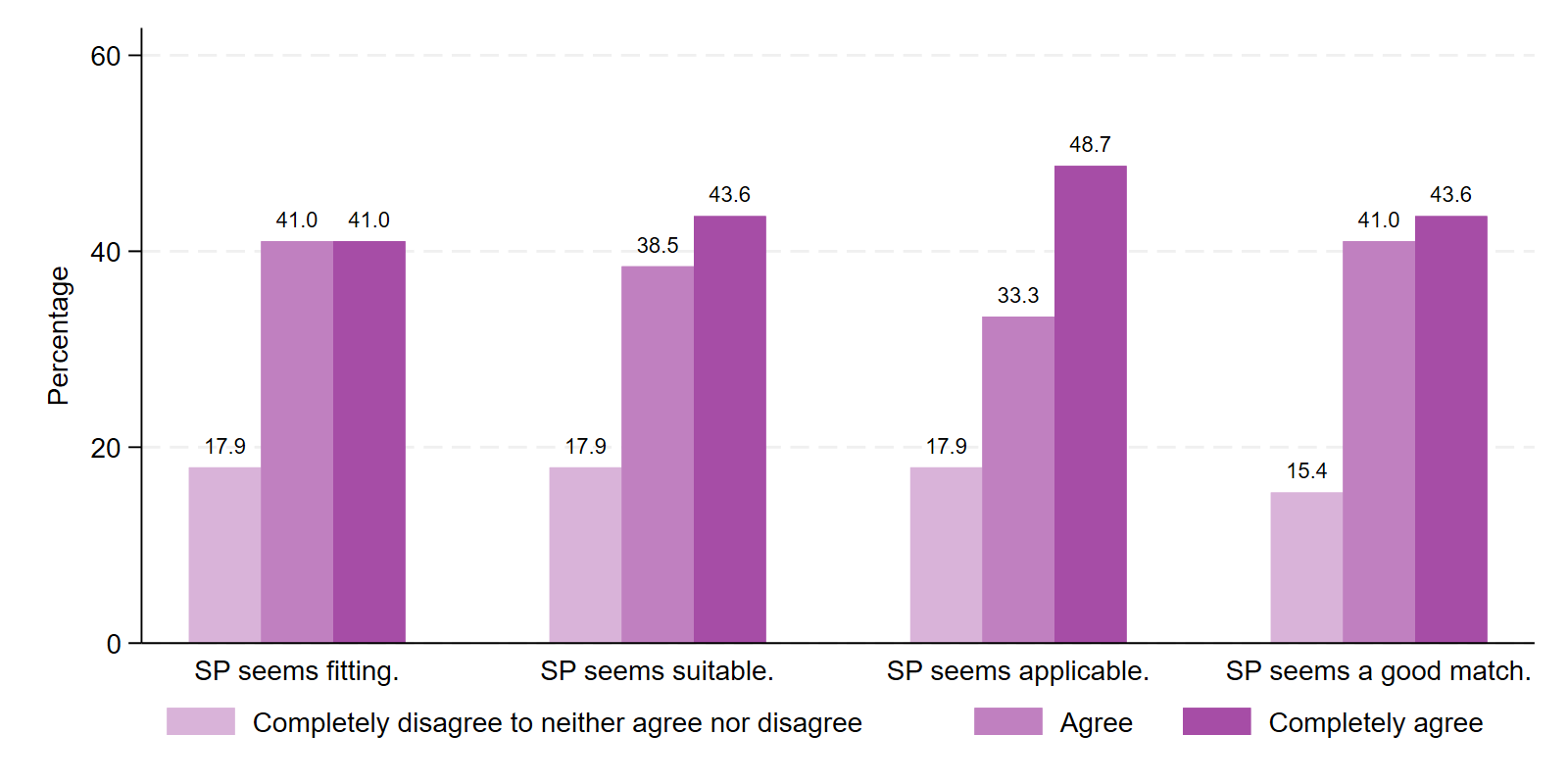

**S6: Table ST1: Results from growth curve models on SDQ total score and subdomains**

|  | SDQ total | | SDQ:  emotional | | SDQ:  conduct | | SDQ:  hyperactive | | SDQ:  peer | | SDQ:  prosocial | |
| --- | --- | --- | --- | --- | --- | --- | --- | --- | --- | --- | --- | --- |
| 3.month | 0.08 | [-0.03,0.18] | -0.07 | [-0.19,0.05] | 0.10 | [-0.01,0.21] | 0.05 | [-0.07,0.16] | 0.13 | [0.01,0.25] | -0.03 | [-0.16,0.09] |
| 6.month | 0.01 | [-0.10,0.13] | -0.20 | [-0.33,-0.07] | 0.04 | [-0.08,0.16] | 0.05 | [-0.07,0.17] | 0.14 | [0.01,0.27] | -0.13 | [-0.26,0.01] |
| SP | 0.13 | [-0.05,0.32] | -0.13 | [-0.31,0.05] | 0.07 | [-0.12,0.26] | 0.19 | [0.00,0.38] | 0.21 | [0.03,0.40] | 0.09 | [-0.09,0.28] |
| 3.month#SP | -0.13 | [-0.28,0.02] | -0.08 | [-0.25,0.08] | -0.12 | [-0.27,0.04] | -0.06 | [-0.22,0.09] | -0.09 | [-0.25,0.07] | 0.04 | [-0.13,0.21] |
| 6.month#SP | -0.16 | [-0.32,-0.00] | 0.07 | [-0.10,0.25] | -0.17 | [-0.34,-0.01] | -0.16 | [-0.33,0.01] | -0.16 | [-0.33,0.01] | 0.20 | [0.02,0.38] |
| Age | 0.03 | [-0.02,0.08] | 0.08 | [0.04,0.13] | -0.02 | [-0.07,0.03] | -0.03 | [-0.08,0.02] | 0.06 | [0.02,0.11] | -0.01 | [-0.05,0.04] |
| Gender: female | 0.27 | [0.08,0.45] | 0.57 | [0.40,0.74] | -0.04 | [-0.23,0.15] | 0.05 | [-0.13,0.24] | 0.13 | [-0.05,0.31] | 0.22 | [0.05,0.40] |
| Gender: other | 0.50 | [-0.07,1.06] | 0.33 | [-0.20,0.86] | 0.34 | [-0.23,0.92] | 0.23 | [-0.34,0.80] | 0.39 | [-0.17,0.94] | -0.00 | [-0.55,0.54] |
| Ethnic minority | 0.01 | [-0.22,0.24] | -0.17 | [-0.38,0.04] | 0.21 | [-0.02,0.44] | 0.02 | [-0.21,0.25] | -0.02 | [-0.25,0.20] | -0.20 | [-0.42,0.03] |
| No school meals | -0.16 | [-0.35,0.04] | -0.06 | [-0.25,0.12] | 0.01 | [-0.18,0.21] | -0.18 | [-0.38,0.01] | -0.18 | [-0.37,0.02] | 0.15 | [-0.04,0.33] |
| IMD 2 | 0.03 | [-0.21,0.26] | 0.16 | [-0.06,0.38] | -0.06 | [-0.30,0.17] | -0.13 | [-0.36,0.11] | 0.10 | [-0.13,0.33] | 0.03 | [-0.20,0.25] |
| IMD 3 | 0.02 | [-0.23,0.27] | 0.10 | [-0.14,0.33] | -0.10 | [-0.36,0.15] | 0.00 | [-0.25,0.25] | 0.05 | [-0.20,0.29] | 0.07 | [-0.18,0.31] |
| IMD 4 | 0.01 | [-0.26,0.29] | 0.18 | [-0.07,0.44] | -0.23 | [-0.51,0.05] | -0.05 | [-0.32,0.23] | 0.13 | [-0.14,0.40] | 0.08 | [-0.19,0.34] |
| IMD 5 | -0.40 | [-0.74,-0.05] | -0.07 | [-0.39,0.25] | -0.49 | [-0.84,-0.14] | -0.26 | [-0.60,0.08] | -0.24 | [-0.58,0.10] | 0.27 | [-0.06,0.60] |
| _cons | -0.21 | [-0.51,0.09] | -0.51 | [-0.79,-0.23] | 0.15 | [-0.15,0.45] | 0.15 | [-0.15,0.45] | -0.35 | [-0.65,-0.06] | -0.30 | [-0.59,-0.01] |
| ln_SD (_cons) | -0.17 | [-0.25,-0.09] | -0.28 | [-0.36,-0.20] | -0.16 | [-0.24,-0.08] | -0.18 | [-0.26,-0.10] | -0.21 | [-0.29,-0.13] | -0.24 | [-0.32,-0.16] |
| ln_SD (residual) | -0.69 | [-0.74,-0.63] | -0.57 | [-0.62,-0.51] | -0.65 | [-0.70,-0.59] | -0.62 | [-0.68,-0.57] | -0.59 | [-0.65,-0.54] | -0.54 | [-0.59,-0.48] |
| n (N) | 448 (1090) | | 448 (1090) | | 448 (1090) | | 448 (1090) | | 448 (1090) | | 448 (1090) | |

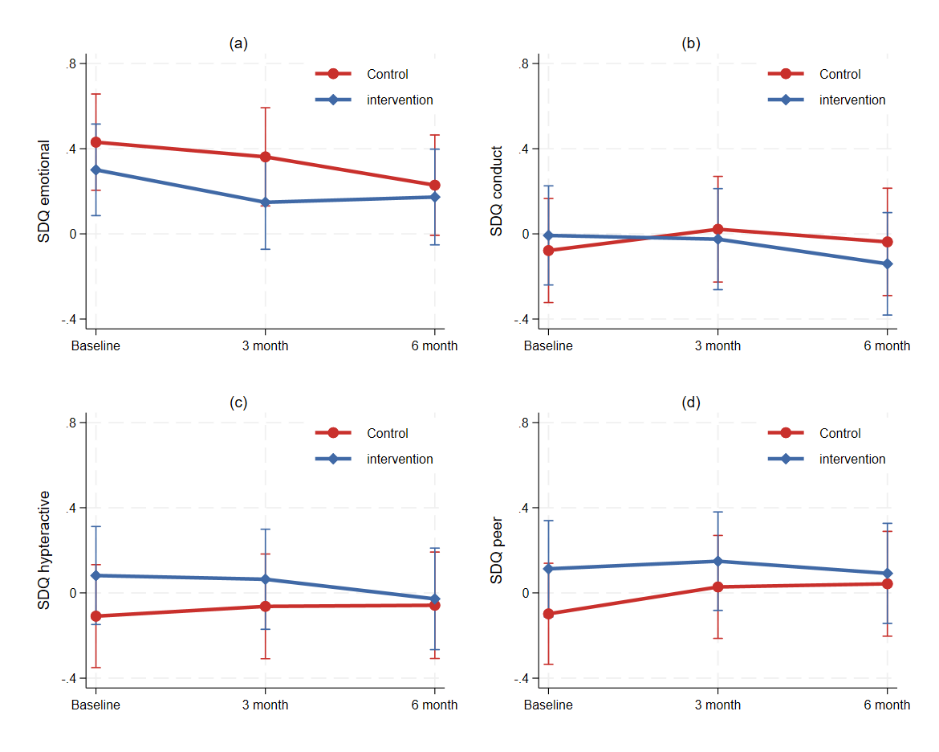
**S7: Figure SF2. Estimated SDQ subdomains across different time points by intervention group**

**S8: Table ST2: Results from growth curve models on PSS, RCADS total score and subdomains**

|  | PSS | | RCADS total | | RCADS:  anxiety | | RCADS:  depression | |
| --- | --- | --- | --- | --- | --- | --- | --- | --- |
| 3.month | -0.21 | [-0.36,-0.06] | -0.06 | [-0.17,0.06] | -0.06 | [-0.18,0.07] | -0.05 | [-0.16,0.07] |
| 6.month | -0.27 | [-0.43,-0.11] | -0.10 | [-0.22,0.02] | -0.11 | [-0.24,0.02] | -0.07 | [-0.19,0.06] |
| SP | -0.16 | [-0.34,0.02] | -0.14 | [-0.31,0.04] | -0.13 | [-0.31,0.05] | -0.12 | [-0.29,0.05] |
| 3.month#SP | 0.04 | [-0.17,0.24] | -0.01 | [-0.17,0.15] | -0.01 | [-0.18,0.15] | -0.01 | [-0.17,0.15] |
| 6.month#SP | 0.05 | [-0.17,0.27] | 0.03 | [-0.13,0.20] | 0.05 | [-0.13,0.23] | 0.01 | [-0.16,0.18] |
| Age | 0.08 | [0.04,0.12] | 0.14 | [0.09,0.18] | 0.08 | [0.03,0.12] | 0.18 | [0.14,0.23] |
| Gender: female | 0.27 | [0.11,0.44] | 0.57 | [0.40,0.75] | 0.55 | [0.37,0.72] | 0.49 | [0.33,0.66] |
| Gender: other | 0.32 | [-0.17,0.82] | 0.60 | [0.08,1.12] | 0.49 | [-0.03,1.02] | 0.61 | [0.10,1.12] |
| Ethnic minority | -0.04 | [-0.24,0.16] | -0.01 | [-0.22,0.20] | -0.16 | [-0.38,0.05] | 0.17 | [-0.03,0.38] |
| No school meals | 0.05 | [-0.11,0.22] | 0.06 | [-0.12,0.24] | 0.10 | [-0.09,0.28] | -0.00 | [-0.18,0.17] |
| IMD 2 | 0.01 | [-0.19,0.22] | 0.12 | [-0.10,0.33] | 0.16 | [-0.06,0.38] | 0.05 | [-0.16,0.26] |
| IMD 3 | 0.11 | [-0.10,0.33] | 0.12 | [-0.11,0.35] | 0.12 | [-0.11,0.36] | 0.09 | [-0.13,0.32] |
| IMD 4 | 0.14 | [-0.10,0.38] | 0.22 | [-0.03,0.47] | 0.16 | [-0.10,0.42] | 0.25 | [0.01,0.50] |
| IMD 5 | -0.03 | [-0.32,0.27] | -0.09 | [-0.41,0.22] | -0.07 | [-0.39,0.25] | -0.10 | [-0.41,0.21] |
| _cons | -0.26 | [-0.53,0.01] | -0.80 | [-1.07,-0.52] | -0.61 | [-0.89,-0.32] | -0.87 | [-1.14,-0.60] |
| ln_SD (_cons) | -0.46 | [-0.56,-0.35] | -0.29 | [-0.37,-0.21] | -0.28 | [-0.36,-0.20] | -0.32 | [-0.41,-0.24] |
| ln_SD (residual) | -0.35 | [-0.40,-0.29] | -0.62 | [-0.67,-0.56] | -0.57 | [-0.62,-0.51] | -0.60 | [-0.66,-0.55] |
| n (N) | 445 (1082) | | 445 (1080) | | 445 (1080) | | 445 (1080) | |

**S9: Figure SF3. Estimated RCADS subdomains across different time points by intervention groups**

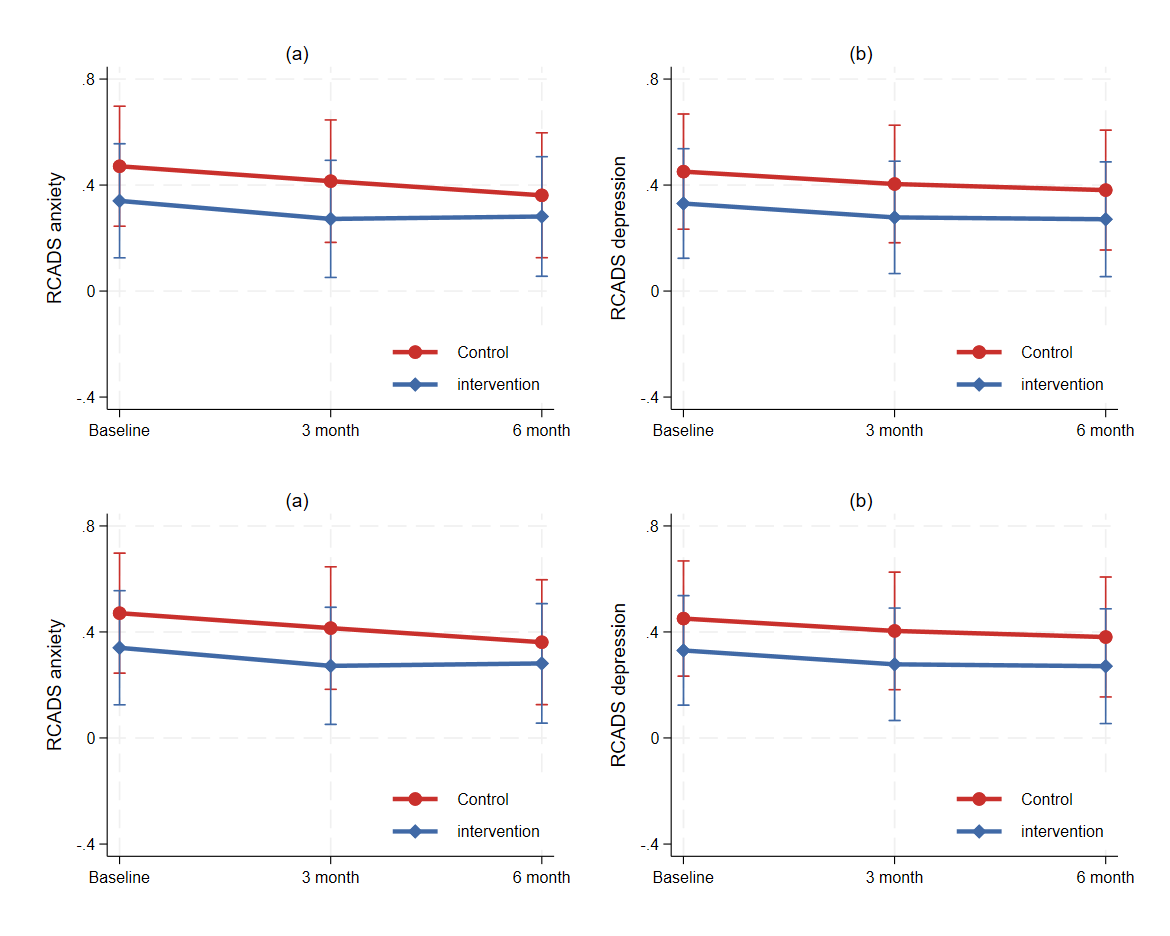

**S10: Table ST3 Results from growth curve models on SRS total score and subdomains**

|  | SRS total | | SRS:  community connection | | SRS:  participation | | SRS:  Self-esteem | | SRS:  empathy | | SRS:  problem solving | | SRS:  goal & aspirations | |
| --- | --- | --- | --- | --- | --- | --- | --- | --- | --- | --- | --- | --- | --- | --- |
| 3.month | 0.06 | [-0.05,0.17] | -0.02 | [-0.15,0.11] | -0.00 | [-0.11,0.11] | 0.15 | [0.02,0.29] | -0.06 | [-0.20,0.07] | 0.11 | [-0.03,0.25] | 0.05 | [-0.08,0.18] |
| 6.month | 0.01 | [-0.11,0.13] | -0.09 | [-0.23,0.04] | -0.09 | [-0.20,0.03] | 0.08 | [-0.06,0.23] | -0.01 | [-0.15,0.13] | 0.11 | [-0.04,0.26] | 0.09 | [-0.04,0.23] |
| SP | -0.02 | [-0.20,0.15] | 0.07 | [-0.12,0.25] | -0.06 | [-0.24,0.12] | -0.06 | [-0.24,0.12] | -0.09 | [-0.27,0.10] | -0.03 | [-0.22,0.15] | 0.06 | [-0.12,0.25] |
| 3.month#SP | 0.04 | [-0.11,0.20] | -0.10 | [-0.28,0.08] | 0.11 | [-0.04,0.26] | 0.00 | [-0.18,0.18] | 0.18 | [-0.01,0.36] | -0.01 | [-0.20,0.18] | 0.07 | [-0.11,0.25] |
| 6.month#SP | 0.23 | [0.07,0.40] | 0.02 | [-0.17,0.21] | 0.23 | [0.07,0.39] | 0.22 | [0.02,0.41] | 0.15 | [-0.05,0.34] | 0.19 | [-0.01,0.39] | 0.09 | [-0.10,0.27] |
| Age | -0.11 | [-0.16,-0.06] | -0.12 | [-0.17,-0.07] | -0.11 | [-0.16,-0.06] | -0.04 | [-0.08,0.01] | 0.04 | [-0.00,0.09] | -0.07 | [-0.11,-0.02] | -0.07 | [-0.12,-0.03] |
| Gender: female | -0.35 | [-0.53,-0.18] | -0.22 | [-0.39,-0.04] | -0.24 | [-0.42,-0.06] | -0.43 | [-0.60,-0.26] | 0.25 | [0.07,0.43] | -0.30 | [-0.47,-0.13] | -0.33 | [-0.50,-0.15] |
| Gender: other | -0.51 | [-1.04,0.03] | -0.79 | [-1.32,-0.26] | 0.01 | [-0.53,0.55] | -0.43 | [-0.95,0.09] | 0.03 | [-0.51,0.57] | -0.16 | [-0.68,0.36] | -0.33 | [-0.87,0.20] |
| Ethnic minority | -0.12 | [-0.34,0.09] | -0.33 | [-0.55,-0.11] | 0.10 | [-0.12,0.32] | 0.05 | [-0.17,0.26] | -0.07 | [-0.29,0.15] | -0.18 | [-0.39,0.03] | 0.06 | [-0.16,0.27] |
| No school meals | 0.13 | [-0.06,0.31] | -0.03 | [-0.21,0.16] | 0.19 | [-0.00,0.37] | 0.11 | [-0.07,0.29] | 0.18 | [-0.01,0.37] | 0.04 | [-0.14,0.22] | 0.02 | [-0.16,0.21] |
| IMD 2 | 0.11 | [-0.11,0.33] | 0.22 | [-0.00,0.44] | 0.03 | [-0.19,0.26] | 0.11 | [-0.11,0.33] | 0.13 | [-0.10,0.35] | -0.06 | [-0.28,0.16] | -0.03 | [-0.25,0.20] |
| IMD 3 | 0.02 | [-0.22,0.26] | 0.08 | [-0.16,0.31] | 0.11 | [-0.13,0.35] | -0.02 | [-0.25,0.21] | 0.14 | [-0.10,0.38] | -0.17 | [-0.40,0.06] | -0.06 | [-0.30,0.18] |
| IMD 4 | 0.00 | [-0.26,0.26] | 0.03 | [-0.23,0.29] | 0.29 | [0.03,0.55] | -0.07 | [-0.32,0.19] | 0.07 | [-0.19,0.33] | -0.21 | [-0.47,0.04] | -0.11 | [-0.37,0.15] |
| IMD 5 | 0.30 | [-0.02,0.62] | 0.16 | [-0.17,0.48] | 0.36 | [0.03,0.68] | 0.19 | [-0.13,0.50] | 0.12 | [-0.21,0.44] | 0.17 | [-0.15,0.48] | 0.16 | [-0.17,0.48] |
| _cons | 0.38 | [0.10,0.67] | 0.50 | [0.22,0.79] | 0.20 | [-0.08,0.49] | 0.22 | [-0.06,0.50] | -0.48 | [-0.77,-0.19] | 0.42 | [0.14,0.70] | 0.33 | [0.05,0.62] |
| ln_SD (_cons) | -0.25 | [-0.33,-0.17] | -0.29 | [-0.38,-0.20] | -0.23 | [-0.31,-0.15] | -0.32 | [-0.41,-0.23] | -0.28 | [-0.37,-0.19] | -0.34 | [-0.44,-0.25] | -0.28 | [-0.37,-0.20] |
| ln_SD (residual) | -0.65 | [-0.71,-0.60] | -0.50 | [-0.56,-0.45] | -0.67 | [-0.73,-0.62] | -0.48 | [-0.53,-0.42] | -0.47 | [-0.52,-0.41] | -0.42 | [-0.48,-0.37] | -0.50 | [-0.56,-0.45] |
| n (N) | 444 (1,078) | | 444 (1,078) | | 444 (1,078) | | 444 (1,078) | | 444 (1,078) | | 444 (1,078) | | 444 (1,078) | |

**S11: Figure SF4. Estimated SRS subdomains across different time points by intervention groups**

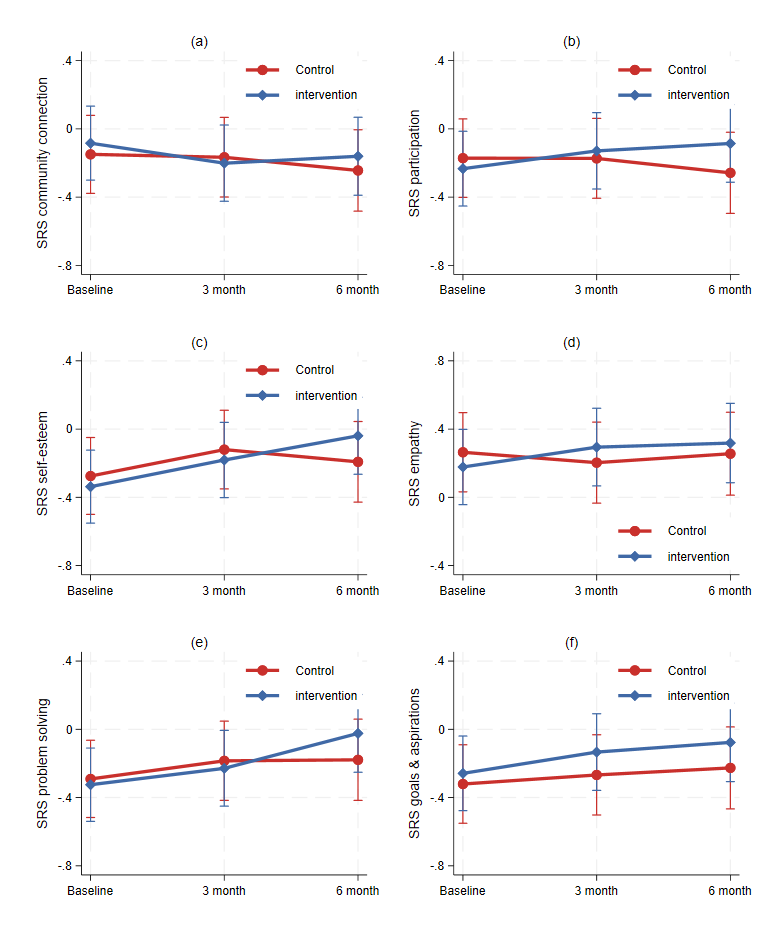

**S12: Table ST4 Results from growth curve models on ONS wellbeing measures**

|  | ONS: life satisfaction | | ONS: worthwhile | | ONS: happiness | |
| --- | --- | --- | --- | --- | --- | --- |
| 3.month | 0.10 | [-0.04,0.23] | 0.02 | [-0.12,0.17] | 0.05 | [-0.11,0.20] |
| 6.month | 0.16 | [0.01,0.30] | 0.12 | [-0.03,0.27] | 0.04 | [-0.12,0.21] |
| SP | 0.08 | [-0.10,0.26] | 0.04 | [-0.14,0.22] | 0.06 | [-0.12,0.24] |
| 3.month#SP | 0.02 | [-0.16,0.21] | 0.10 | [-0.09,0.29] | -0.00 | [-0.21,0.21] |
| 6.month#SP | 0.05 | [-0.14,0.25] | 0.08 | [-0.12,0.28] | 0.14 | [-0.08,0.36] |
| Age | -0.13 | [-0.17,-0.09] | -0.13 | [-0.17,-0.08] | -0.11 | [-0.15,-0.06] |
| Gender: female | -0.46 | [-0.63,-0.30] | -0.33 | [-0.50,-0.16] | -0.36 | [-0.53,-0.20] |
| Gender: other | -0.64 | [-1.15,-0.13] | -0.73 | [-1.25,-0.22] | -0.69 | [-1.18,-0.19] |
| Ethnic minority | -0.10 | [-0.31,0.11] | -0.08 | [-0.29,0.13] | -0.12 | [-0.33,0.08] |
| No school meals | -0.04 | [-0.22,0.14] | -0.05 | [-0.23,0.12] | -0.03 | [-0.20,0.14] |
| IMD 2 | 0.09 | [-0.13,0.30] | -0.07 | [-0.28,0.15] | -0.04 | [-0.25,0.16] |
| IMD 3 | -0.15 | [-0.38,0.07] | -0.11 | [-0.34,0.11] | -0.16 | [-0.38,0.06] |
| IMD 4 | -0.23 | [-0.48,0.02] | -0.12 | [-0.37,0.13] | -0.31 | [-0.55,-0.07] |
| IMD 5 | -0.08 | [-0.39,0.23] | -0.07 | [-0.38,0.24] | -0.12 | [-0.42,0.18] |
| _cons | 0.68 | [0.40,0.95] | 0.61 | [0.33,0.88] | 0.64 | [0.38,0.91] |
| ln_SD (_cons) | -0.35 | [-0.44,-0.26] | -0.37 | [-0.46,-0.27] | -0.46 | [-0.57,-0.36] |
| ln_SD (residual) | -0.47 | [-0.52,-0.41] | -0.41 | [-0.47,-0.36] | -0.34 | [-0.39,-0.29] |
| n (N) | 445 (1080) | | 445 (1080) | | 445 (1080) | |
